# Robust AI Framework for Comprehensive Tuberculosis Drug Resistance Profiling with Rapid Adaptability

**DOI:** 10.64898/2026.07.20.26358475

**Authors:** Chang Liu, Haoyue Zhu, Xingze Wang, Yan Li, Hui Wang, Yang Yang

## Abstract

Tuberculosis remains the leading cause of death from a single infectious agent, with drug-resistant tuberculosis, particularly multidrug-resistant and extensively drug-resistant strains, posing major challenges for timely treatment. Whole-genome sequencing can accelerate resistance detection, but current genomic and machine-learning approaches typically predict resistance to individual drugs, do not directly infer regimen-relevant resistance profiles, and generalise poorly across regions or newly introduced drugs. We developed *MuseAMR*, a multimodal, multi-label deep-learning framework that predicts both individual-drug resistance and clinically actionable composite phenotypes from *Mycobacterium tuberculosis* genomes, with robust cross-regional performance and few-shot adaptation to emerging drugs. Trained on 10,886 isolates and externally validated on 18,334 isolates from six global regions, MuseAMR improved sensitivity for second-line drug resistance (0.857 versus 0.655) and MDR/pre-XDR profiles compared with WHO catalogue-based prediction while maintaining high specificity. It also showed robust cross-regional performance and few-shot adaptation to bedaquiline, delamanid and linezolid using 5-20 resistant isolates, with attribution analyses recovering established resistance loci. These results support its potential for regimen-level tuberculosis resistance profiling and surveillance.

## Introduction

Tuberculosis (TB), caused by Mycobacterium tuberculosis (MTB), is the leading cause of death from a single infectious agent worldwide, and drug-resistant strains drive disproportionately high mortality ^1,2^. Despite being treatable, drug-resistant TB (DR-TB) is associated with prolonged treatment, high relapse risk, extensive antibiotic exposure, and worse outcomes, especially in patients with chronic comorbidities ^3^. The clinical gold standard for determining drug resistance is the culture-based phenotypic drug susceptibility test (pDST), which can take up to 8 weeks. To combat DR-TB, the World Health Organization (WHO) urgently calls for rapid and comprehensive drug susceptibility testing ^1,4,5^. Genotypic drug susceptibility testing (gDST) based on whole-genome sequencing (WGS) offers a much faster alternative, providing high-resolution coverage of the MTB genome ^6,7^. However, identifying resistance-conferring mutations from WGS remains laborious, requiring large datasets, statistical association, and ongoing curation.

Catalogued mutations, such as those in the WHO mutation catalogue, are used clinically ^8–10^. Nevertheless, they miss rare or novel mutations and fail to detect low-abundance hetero-resistance, leading to false-susceptible results. Moreover, they provide no information on newer agents or repurposed drugs, limiting their utility for guiding complex regimens, and maintaining the catalogue demands substantial effort, especially during a pandemic (Figure 1A).

**Figure 1.**
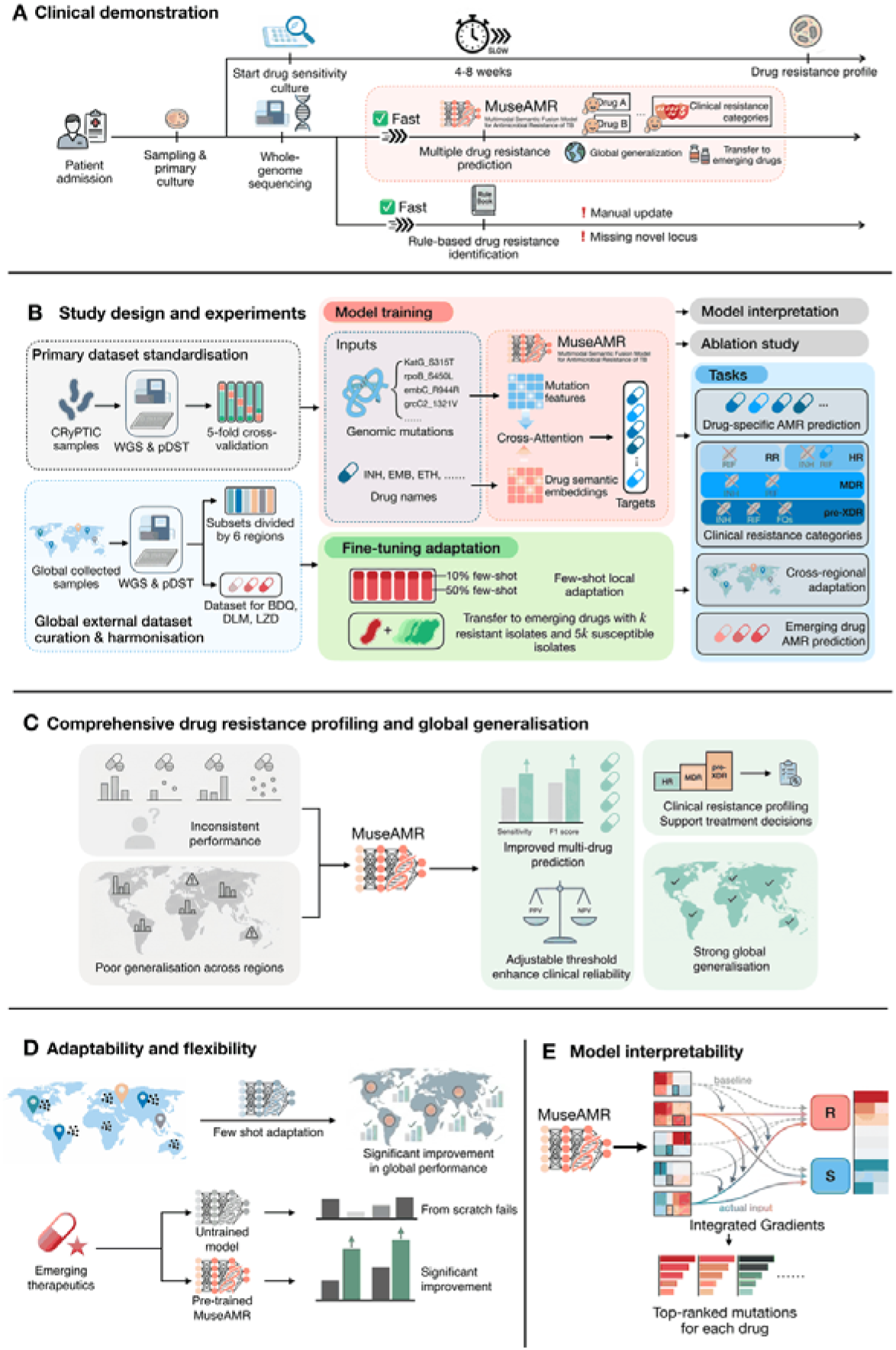
Overall framework and integrated capabilities of MuseAMR for MTB drug resistance profiling. **(A)** Clinical demonstration and implementation scenario. Current workflows take up to 8 weeks for drug susceptibility culture after sampling. In contrast, the proposed MuseAMR pipeline utilises whole-genome sequencing to enable fast, simultaneous resistance prediction for multiple drugs, along with global applicability, clinical resistance categorisation, and transferability to emerging agents. Existing rule-based methods, however, rely on routine manual updates and therefore miss novel resistance loci. **(B)** Integrated study design and experimental workflow. Diagram integrates model architecture, data partitioning, evaluation schemes, and adaptation strategies. The MuseAMR architecture fuses high-dimensional genomic mutation features (processed by a CNN backbone) and low-dimensional drug semantic embeddings using a cross-attention mechanism for multi-label resistance prediction. Model training and validation utilised paired WGS and phenotypic data: a primary development dataset for 5-fold cross-validation, and global external validation cohorts stratified into six regions. Evaluation included drug-specific performance, aggregate metrics, clinical resistance categories, and ablation studies. To address regional distribution shifts, few-shot fine-tuning adaptation strategies were developed using *k* positive and 5*k* negative samples at two data proportions (10% and 50% few-shot samples). For emerging drugs (BDQ, DLM, LZD), the same adaptation protocol was applied. **(C)** Comprehensive drug resistance profiling and global generalisation. Compared to existing methods, MuseAMR overcomes inconsistent performance across drugs and poor regional generalisation, delivering improved multi-drug prediction (high sensitivity and F1-score), clinical resistance profiling from HR to XDR to guide treatment decisions, and robust global generalisation. Additionally, its adjustable thresholds enhance clinical reliability by optimising positive and negative predictive values (PPV and NPV). **(D)** Model adaptability and flexibility. Model performance is enhanced through adaptation strategies. (Top) Few-shot adaptation significantly boosts global performance on region-specific datasets. (Bottom) For emerging therapeutics (e.g., BDQ), training from scratch fails, whereas fine-tuning the pretrained MuseAMR achieves substantial performance gains with minimal data. **(E)** Model interpretability. Diagram shows the identification of key features driving drug-specific predictions. Using Integrated Gradients to compare actual inputs against a baseline, the model highlights and ranks the top genomic mutations that are critical for either resistant (R) or sensitive (S) classifications.

Artificial intelligence (AI) offers a promising complementary approach. By learning complex patterns from large-scale genomic and pDST data, AI models can predict resistance to multiple drugs ^11–14^. However, existing models typically operate on a drug-by-drug basis, resulting in uneven performance: strong for first-line agents such as rifampicin and isoniazid, but weaker for second-line and emerging drugs ^11,14^. This inconsistency is clinically problematic because WHO treatment pathways are organised around bundled resistance categories, such as isoniazid-resistant (HR) TB, multidrug / rifampicin-resistant (MDR/RR) TB, and pre-extensively drug-resistant (pre-XDR) TB, rather than individual single-drug predictions ^15^. A patchwork of drug-specific models with variable reliability cannot support dependable regimen-level decision support.

Beyond per-drug inconsistency, existing WGS-based AI models suffer from poor generalisability across epidemiological contexts and over time. They often show marked performance declines in discrimination when applied to geographically distinct cohorts, different lineage structures, or drugs with few resistant isolates in the training set ^14,16^. A global benchmark study confirmed persistent trade-offs between discrimination, calibration, and drug coverage, with many approaches remaining constrained for less well-represented drugs ^11^. Thus, clinically deployable systems must shift from static, drug-specific predictors to flexible, extendable architectures that can accommodate evolving resistance landscapes without complete retraining.

To address these gaps, we developed the Multimodal Semantic Fusion Model for Antimicrobial Resistance (MuseAMR), an end-to-end multimodal, multi-label deep learning framework. Unlike previous drug-by-drug methods, MuseAMR directly profiles composite resistance phenotypes from WGS data, jointly predicting resistance to nine anti-tuberculosis drugs and classifying clinically actionable resistance categories aligned with WHO care pathways (e.g., MDR/RR-TB, pre-XDR-TB). We conducted large-scale multinational external validation across six global cohorts and demonstrated robust cross-regional generalisation with few-shot local recalibration. Crucially, using only 5-20 resistant isolates, the model adapts rapidly to emerging drugs (bedaquiline, delamanid, linezolid) via few-shot transfer learning, without requiring any chemical or structural information. This dual capacity for generalisation and adaptation directly addresses the limitations of current approaches and provides a robust tool for precision TB management and resistance surveillance.

## Results

### Dataset characteristics and resistance profiles

Following quality control and phenotypic standardisation, the primary CRyPTIC-derived dataset comprised 10,886 Mycobacterium tuberculosis isolates from 9 countries/regions. For external validation, we curated 18,334 isolates from 25 independent studies and stratified them into 6 regional cohorts: China, Africa, Oceania, Asia excluding China, the Americas, and Europe (Figure 2A-B). China accounted for 47.2% of the external validation dataset and was analysed separately to avoid overrepresenting one high-sample region in cross-regional estimates.

**Figure 2.**
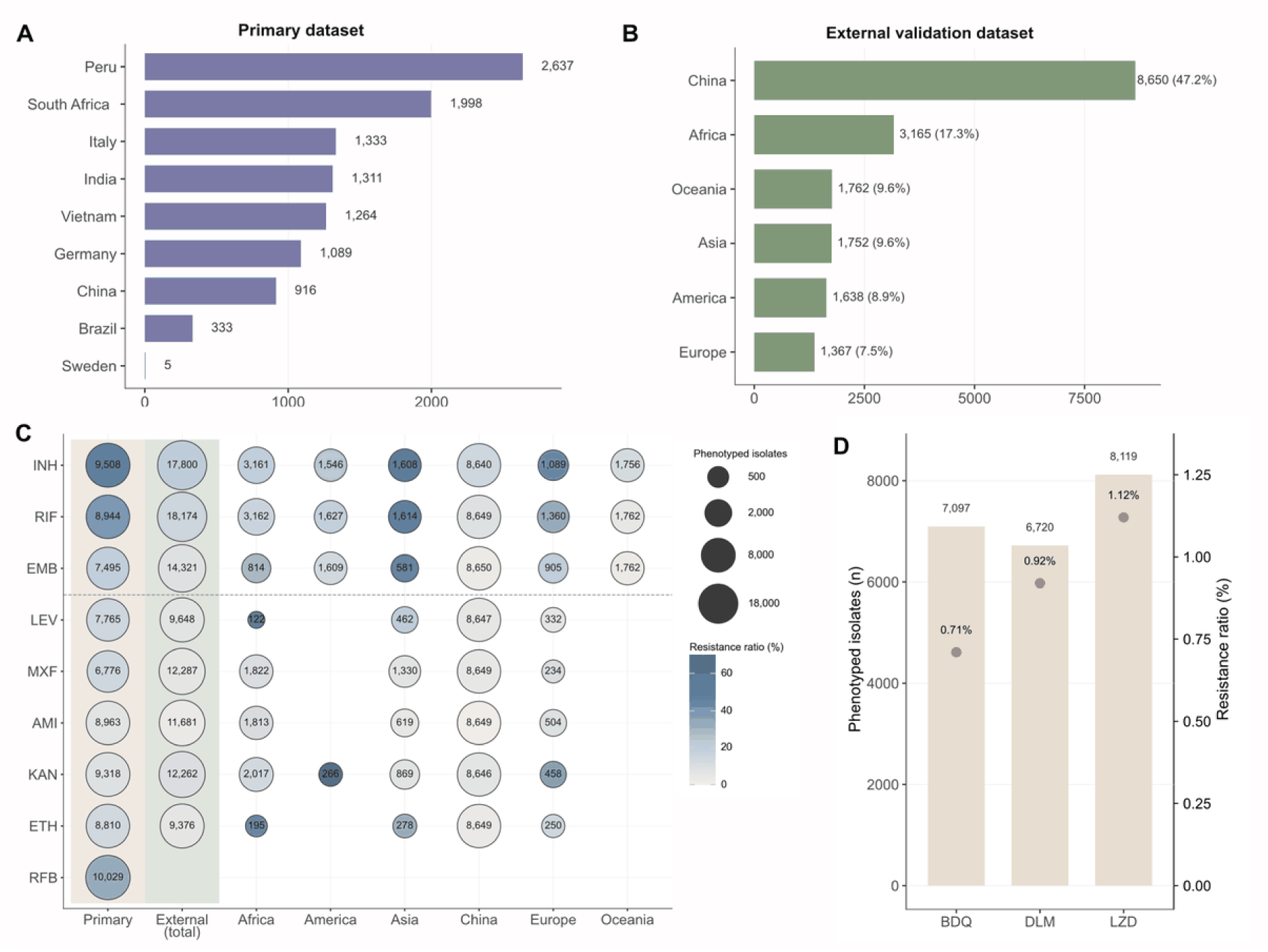
Overview of composition and resistance landscape across the study datasets. The figure summarises the composition and resistance landscape distribution of the primary dataset, the external validation dataset, and the emerging drug extension dataset. **(A)** The number of isolates from each country in the primary dataset; **(B)** The external validation dataset is divided into six subsets according to the sample origins, and the bar chart shows the sample count and proportion of each region in the validation set; **(C)** The number of phenotyped isolates and resistance ratios in each dataset (subset), where larger bubbles indicate more phenotyped isolates, and darker colours indicate higher resistance ratios; **(D)** The number of phenotyped isolates and resistance proportions of three constructed new drugs in the additionally collected emerging drug extension dataset, with bars representing the number of phenotyped isolates (left axis) and points representing the resistance ratios (right axis).

The main modelling analysis included three first-line drugs (INH, RIF, and EMB) and six second-line drugs (LEV, MXF, AMI, KAN, ETH, and RFB). Phenotypic availability was highest for first-line drugs and more uneven for second-line drugs; notably, RFB phenotypes were unavailable in all six external cohorts (Figure 2C). Resistance proportions also varied across datasets, with generally higher resistance in the primary dataset.

For the emerging drug extension analysis, we aggregated isolates with phenotypic records for BDQ, DLM, and LZD. Resistance was rare for all three drugs, ranging from 0.71% for BDQ to 1.12% for LZD, creating a highly imbalanced few-shot transfer learning setting (Figure 2D).

### MuseAMR outperforms existing methods in predicting clinical resistance categories

On the primary dataset, we first compared MuseAMR with WDNN and LR, using 5-fold cross-validation. For first-line drugs, MuseAMR achieved the best overall balance across the aggregated metrics. Its sensitivity reached 0.93, while specificity remained 0.93 (Figure 3A). For second-line drugs, MuseAMR (PE) achieved a sensitivity of 0.854, exceeding WDNN (0.830, *p*=0.07) and LR (0.826, *p*<0.05). The main advantage of MuseAMR is a more favourable diagnostic trade-off for the more challenging second-line tasks.

**Figure 3.**
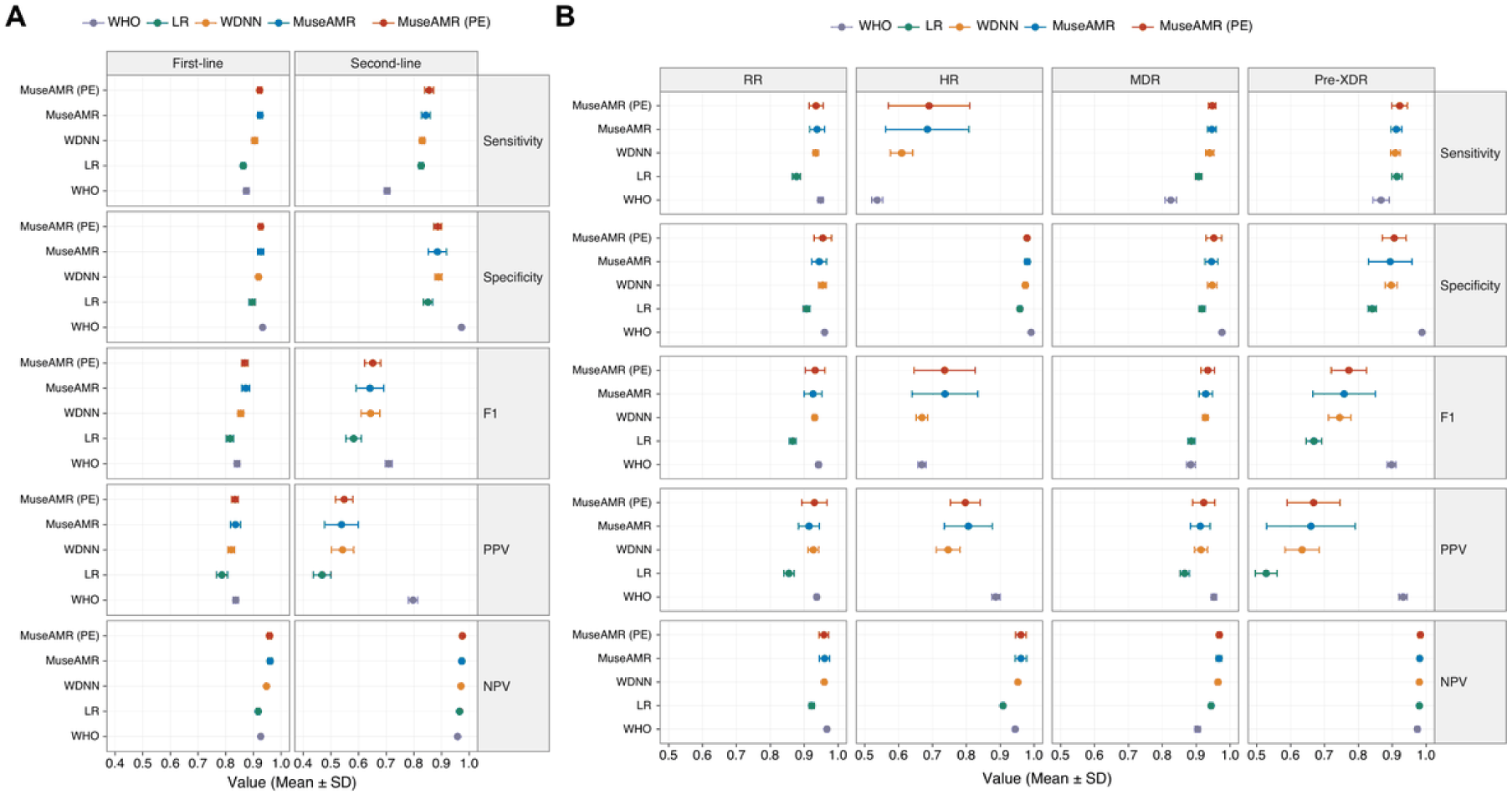
Predictive performance of MuseAMR for individual drug classes and WHO-defined clinical resistance categories in internal cross-validation. **(A)** Average performance metrics across first-line and second-line anti-tuberculosis drugs. Five models (MuseAMR with and without protein embeddings (PE), WDNN, LR, and the WHO catalogue baseline) are compared across Sensitivity, Specificity, F1 score, PPV, and NPV. **(B)** Predictive performance for WHO-defined composite clinical classifications, encompassing Rifampicin-resistant (RR), Isoniazid-resistant (HR), Multidrug-resistant (MDR), and Pre-extensively drug-resistant (Pre-XDR) tuberculosis. In both panels, point estimates denote the mean performance across k-fold cross-validation, with error bars representing the standard deviation (Mean +/- SD). Of note, Sensitivity, F1 score, and PPV for the LR model in the HR classification task are absent from the plot, as their values fall below the lower bound of the displayed x-axis range.

This advantage extended to WHO-aligned clinical resistance profiling (Figure 3B). For HR, MDR, and pre-XDR classifications, MuseAMR consistently improved sensitivity and F1 over baselines, with the largest gains in HR (sensitivity 0.690 vs. 0.610 for WDNN, *p*=0.34; vs. 0.27 for LR, *p*<0.05) and pre-XDR (F1 0.772 vs. 0.744 for WDNN, *p*=0.5; vs. 0.669 for LR, *p*<0.05). RR was already well classified by all learned models, leaving limited room for improvement. Compared with the WHO Catalogue, MuseAMR increased sensitivity for second-line drugs from 0.655 to 0.857, while maintaining specificity. For first-line drugs, MuseAMR also improved sensitivity (0.926 vs. 0.873).

### MuseAMR achieves robust zero-shot cross-regional generalisation

We next evaluated zero-shot transferability across geographically distinct populations without local retraining (Figure 4).

**Figure 4.**
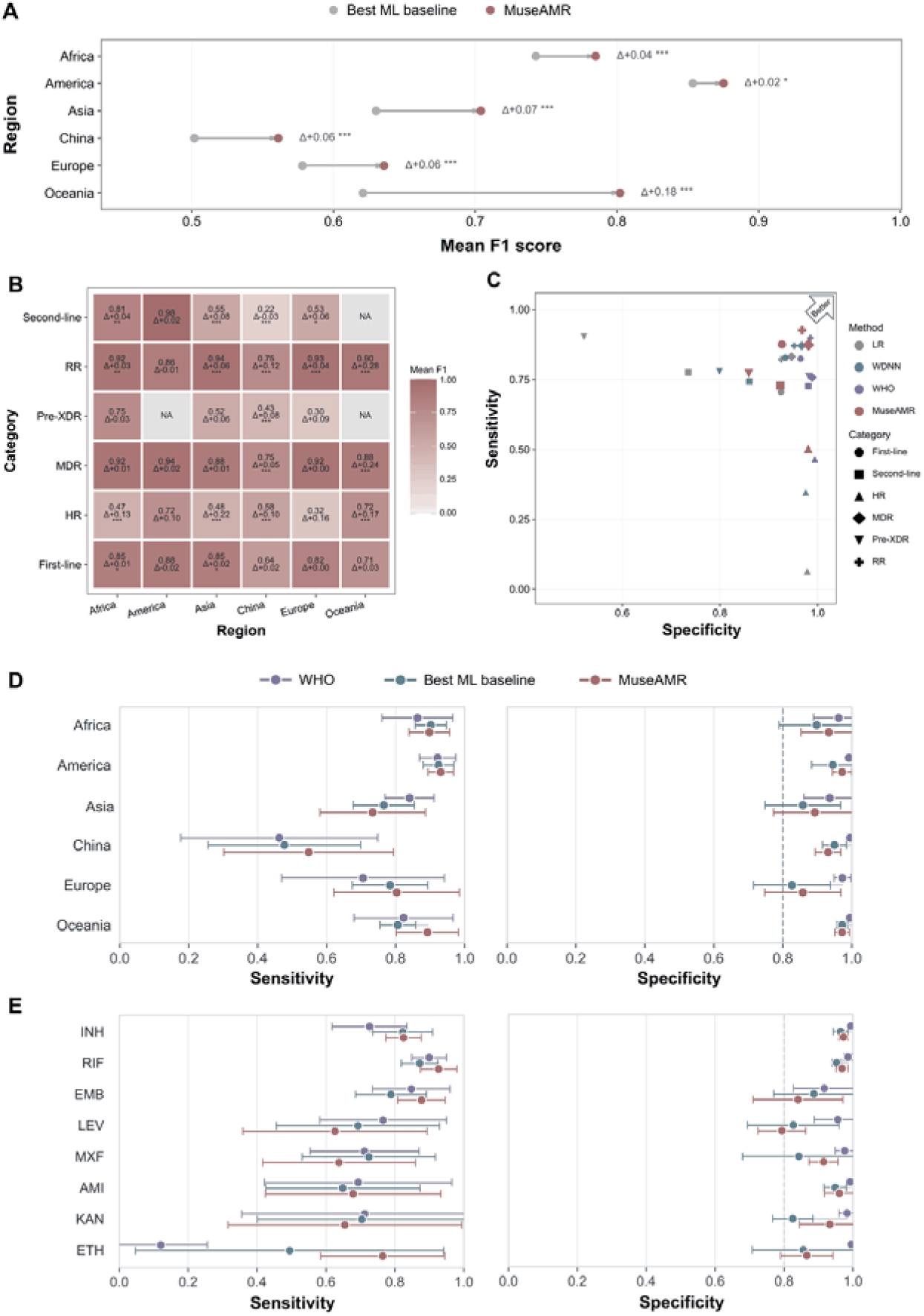
Robust cross-regional generalisation and diagnostic performance of MuseAMR across global cohorts. **(A)** Dumbbell plot illustrating the average performance improvement of MuseAMR over the best-performing ML baselines. Dots represent the mean F1 score across all tasks for each distinct geographic region, with connecting lines and annotations (Delta) denoting the absolute performance gain. **(B)** Heatmap detailing the region-specific mean F1 scores for both individual drug classes (First-line, Second-line) and clinical categories (HR, MDR, Pre-XDR, RR). Each cell displays the absolute F1 score of MuseAMR and its improvement (Delta) relative to the maximal ML baseline. Grey cells indicate missing data (NA) due to insufficient sample sizes. **(C)** Scatter plot evaluating the diagnostic trade-off between Sensitivity (y-axis) and Specificity (x-axis). Each point represents a specific category’s performance aggregated globally, with shapes distinguishing the clinical/drug categories and colours differentiating the predictive methods (LR, WDNN, WHO, MuseAMR). Positions closer to the upper-right corner indicate superior classification reliability. **(D-E)** Point-range plots comparing the Sensitivity (left panels) and Specificity (right panels) among WHO guidelines, the Best ML baseline, and MuseAMR. Performance is evaluated across different geographic regions (D) and across individual anti-tuberculosis drugs (E). Points represent the mean values, and error bars indicate the standard deviations. *, **, and *** denote statistical significance at *p* < 0.05, *p* < 0.01, and *p* < 0.001, respectively.

In zero-shot external validation across six regions, MuseAMR improved the average F1 score over the best ML (LR, WDNN) baseline in each region, ranging from +2% in the Americas to +18% in Oceania (*p*<0.05 for all) (Figure 4A). Mean F1 ranged from 0.876 (America) to 0.562 (China), reflecting varying baseline difficulty across cohorts.

Task-level heterogeneity was evident (Figure 4B): first-line F1 remained above 0.90 in most regions, whereas second-line and composite categories were more variable (e.g., second-line F1 0.30-0.75; MDR F1 0.32-0.58). Notably, the relative gains of MuseAMR were largest for these harder tasks (e.g., MDR +0.22 in Asia excluding China, first-line +0.28 in Oceania, pre-XDR +0.24 in Africa).

The sensitivity-specificity analysis showed that MuseAMR predictions clustered toward the upper-right region of the performance space compared with baselines (Figure 4C). Region-level comparisons showed that MuseAMR achieved comparable or higher sensitivity in Africa, Europe and Oceania, while maintaining high specificity across most regions (Figure 4D). At the drug level, MuseAMR showed improved sensitivity for INH, RIF, EMB, ETH and retained specificity above 0.8 for most drugs (Figure 4E).

### Few-shot adaptation enables rapid regional recalibration and extension to emerging drugs using minimal resistant isolates

For regional adaptation, the pretrained model was fine-tuned with 10% or 50% of local samples and evaluated on the remaining isolates. For emerging drugs, the pretrained backbone was transferred to BDQ, DLM, and LZD using only k=5 or k=20 resistant isolates with matched susceptible samples.

Mean F1 scores across all tasks increased from 10% to 50% fine-tuning (Figure 5A). Larger gains were observed for lower-performing zero-shot tasks. F1 improved by 6% for second-line drugs (0.62 to 0.68, *p*<0.05) and by 7% for pre-XDR (0.50 to 0.57, *p*=0.247). Region-task stratification (Figure 5B) confirmed that harder endpoints yielded larger gains. Representative improvements at 50% fine-tuning include: second-line +12.1 in China (*p*<0.05), pre-XDR +24.7 in Asia excluding China (*p*<0.05), and RR +8.3 in China (*p*<0.05). Gains were smaller where zero-shot performance was already strong (e.g., Americas, Africa). Full region-task results are in Supplementary Table S8.

**Figure 5.**
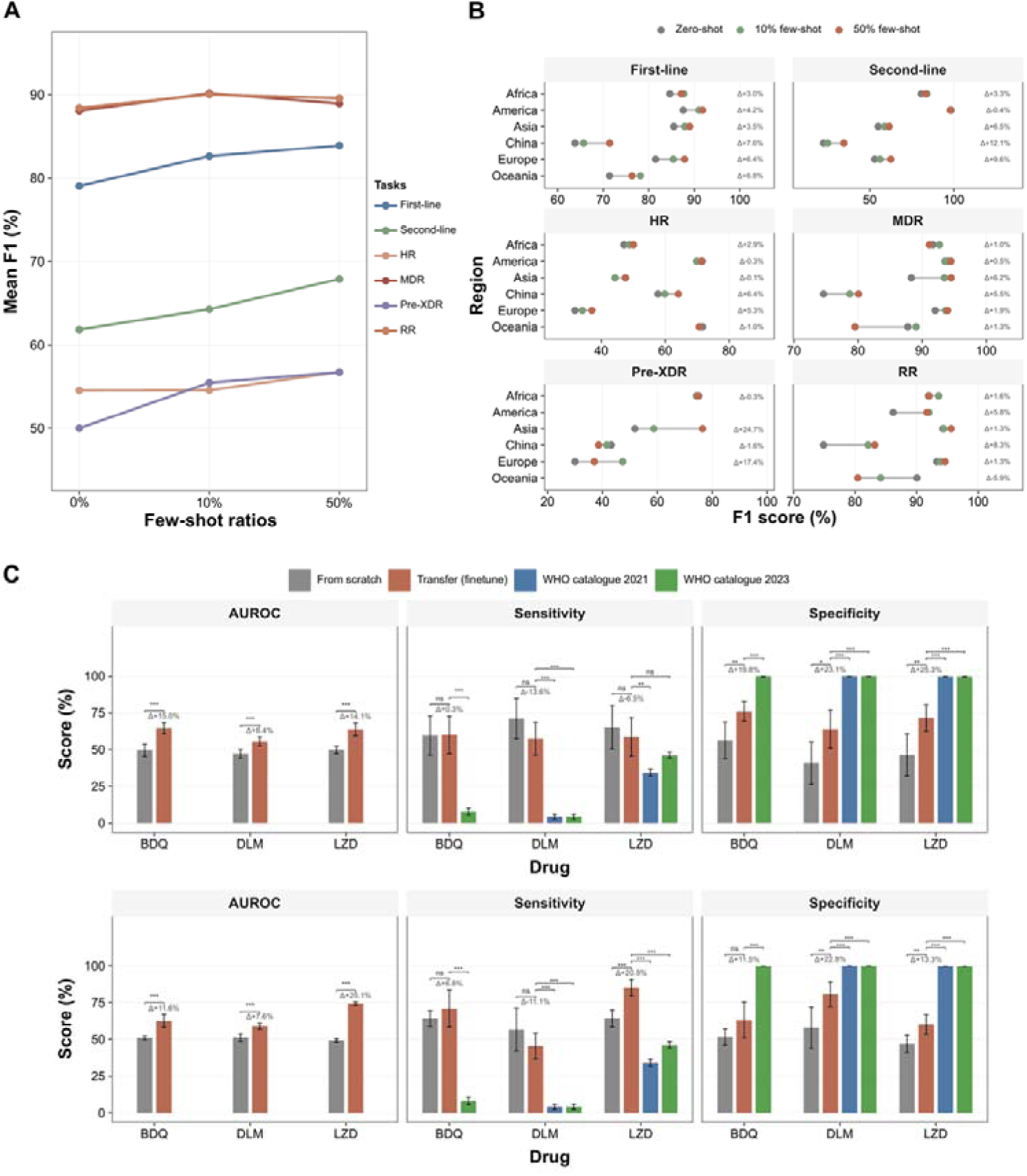
Flexibility, local adaptation, and few-shot transferability of MuseAMR across global cohorts and emerging therapeutics. (**A**) Global learning trajectories illustrating the progressive enhancement of predictive performance (Mean F1 score) with increasing few-shot calibration ratios (0, 10%, 50%). Lines represent the averaged trends across diverse drug classes and clinical categories. **(B)** Regional dumb-bell plots highlighting the local adaptation dynamics of few-shot learning. Points track the transition of F1 scores from the zero-shot baseline (grey) to 10% (green) and 50% (red) local data fine-tuning across six subsets. Numeric annotations indicate the maximum absolute percentage point gain (Delta) achieved during few-shot fine-tuning. **(C)** Transfer learning efficacy for emerging and repurposed anti-tuberculosis drugs (Bedaquiline [BDQ], Delamanid [DLM], and Linezolid [LZD]). Bar plots compare the predictive performance of models trained entirely from scratch (grey) against models leveraging transfer learning (red). The upper and lower rows represent performance at k=5 and k=20 settings, respectively. Evaluated metrics include AUROC, Sensitivity, and Specificity, with corresponding absolute performance shifts (Delta) annotated above the bars. Error bars denote the 95% confidence intervals (CIs) across episodes.

Local calibration also improved diagnostic trade-offs: in China, second-line sensitivity rose from 0.404 to 0.555 (*p*<0.05); in Europe, first-line specificity rose from 0.892 to 0.965 (*p*<0.05). Drug-specific gains (e.g., LEV sensitivity 0.645 to 0.755) are detailed in Supplementary Table S8.

For emerging drugs such as BDQ, DLM, and LZD, we evaluated whether the pre-trained backbone provides transferable knowledge by comparing training from scratch with few-shot transfer learning using *k* = 5 or *k* = 20 resistant isolates matched 1:5 with susceptible isolates (Figure 5C).

From-scratch models achieved AUROCs of about 0.5 both under *k* = 5 and *k* = 20, whereas transfer models reached AUROCs of 0.65, 0.56, and 0.64 for BDQ, DLM, and LZD under k=5 setting (*p*<0.05 for all). Under k=20, the AUROCs of transfer models reached 0.63, 0.59, and 0.74 (*p*<0.05 vs. scratch for all).

Transfer learning also substantially improved specificity (BDQ +0.20, DLM +0.23, LZD +0.25, *k* = 5; all *p*<0.05), while sensitivity remained comparable to scratch baselines. This specificity gain minimises false-positive resistance calls, safeguarding last-resort drugs from being inappropriately withheld. The frozen pretrained representations stabilise learning on micro-scale imbalanced datasets where from-scratch models are dominated by sampling variance. As a rule-based benchmark, the WHO 2023 catalogue achieved high specificity but limited sensitivity for emerging drugs (BDQ 0.08, DLM 0.04, LZD 0.46), all significantly below the transfer models (*p*<0.05 for BDQ and DLM under both *k* settings; LZD *p*<0.05 when *k* = 20).

### Improved model interpretability to enhance clinical trust and readiness

Integrated Gradients attribution showed that MuseAMR recovered established resistance biology despite using whole-genome mutation profiles without predefined gene panels (Supplementary Figure S2). In true-positive isolates, high-attribution features mapped to canonical drug-specific loci, including *katG* for INH, *rpoB* for RIF, *embA*/*embB* for EMB, and *gyrA* for fluoroquinolones.

Misclassified isolates showed more diffuse attribution patterns. False-positive predictions often included canonical resistance mutations together with recurrent background loci, whereas false-negative predictions, particularly for ETH, had broader genomic weighting consistent with heterogeneous resistance mechanisms.

### Architectural ablation reveals semantic integration as core innovation

A systematic ablation analysis (supplementary material) showed that semantic drug embeddings provide drug-conditioned representations, particularly beneficial for second-line drugs. Positional encoding ensures effective capture of genomic spatial information. The gating module does not add discrimination gains but is valuable for stabilising training and efficiency, further demonstrating the superiority of our model’s end-to-end processing of whole-genome features.

## Discussion

The clinical translation of WGS-based ML models for DR-TB remains limited by two interrelated barriers: most existing tools are optimised for isolated drug-level predictions rather than regimen-oriented clinical decisions, and their performance degrades under geographic distribution shifts or when phenotypic data are scarce for less-characterised drugs. In this multinational study, we developed and evaluated MuseAMR using a primary dataset of >10,000 Mycobacterium tuberculosis isolates and an external validation cohort of >18,000 isolates from six global regions. Our findings show that MuseAMR directly addresses both barriers by (1) integrating drug-level predictions into clinically meaningful composite resistance aligned with WHO care pathways; (2) maintaining transportable zero-shot performance across diverse regions; (3) enabling efficient few-shot local recalibration; and (4) supporting rapid extension to emerging drugs using only 5-20 resistant isolates, without requiring any chemical or structural information. Together, these properties move WGS-based DST closer to the requirements of regimen-based care.

Beyond higher per-drug accuracy, MuseAMR produced a more clinically coherent pattern of performance across the resistance spectrum. Gains were concentrated on second-line drugs and on WHO-defined resistance categories (HR-TB, MDR-TB, and pre-XDR-TB), whereas prediction of first-line agents was already strong across methods. This distinction is clinically important because treatment selection for DR-TB is determined by resistance profiles across multiple drugs rather than by isolated resistance calls ^15^. In this context, the value of MuseAMR lies in shifting WGS-based prediction from disconnected single-drug endpoints towards multidrug resistance profiling, particularly in settings where under-detection has the greatest clinical consequence^12^. In the primary dataset, MuseAMR outperformed ML baselines and showed higher sensitivity than the WHO Catalogue. However, because the WHO Catalogue was derived from a large, curated dataset that included isolates overlapping with our primary dataset, this comparison should be interpreted as benchmarking against an expert rule-based reference rather than as a fully independent external comparison.

These findings are consistent with the proposed model architecture. By jointly learning all drug labels while conditioning genomic features on drug semantic embeddings, MuseAMR exploits shared resistance structure, co-resistance patterns, and drug-specific mutational signals simultaneously. This property is particularly valuable for drugs such as ETH and for composite classifications (HR-TB and pre-XDR-TB), where resistance is biologically heterogeneous and catalogue-based approaches often favour specificity over recall ^8,9,11^. The improvement in clinically derived categories therefore suggests that the model is learning resistance relationships at the level at which treatment decisions are made.

The external validation results further strengthen the interpretation that MuseAMR captures transferable resistance structure rather than fitting a single development cohort. Although zero-shot performance remained heterogeneous across the six regions, the model improved mean F1 over the best ML baselines in every external cohort, with gains ranging from +2% in the Americas to +18% in Oceania, despite marked distribution shifts. This is notable because regional transportability and lineage dependency remain persistent barriers in WGS-based resistance prediction ^14,16^.

The additional gains observed after few-shot local recalibration suggest that limited local phenotypic data can offset performance losses caused by inter-regional distribution shifts, particularly in cohorts where zero-shot generalisation was most challenging. Rather than indicating fragility, they support a staged deployment model: a pretrained model developed on multinational data can be recalibrated as local phenotyped isolates accumulate. Such a strategy is especially relevant for TB programmes that cannot assemble very large local training sets at the outset but can progressively improve performance through limited local phenotyping ^17–19^.

The transfer-learning experiments extend the same principle from geographic adaptation to therapeutic evolution. BDQ, DLM, and LZD are clinically important agents in contemporary DR-TB treatment, yet resistant isolates remain scarce for several of these drugs, constraining the development of reliable data-driven predictors ^15^. Under these data-sparse conditions, MuseAMR retained useful discriminative ability and consistently outperformed both de novo training and catalogue-based rules. Notably, this adaptation does not rely on any prior chemical or structural knowledge of the drug; instead, it leverages resistance-associated genomic patterns learned from established drugs, transferred via few-shot learning from only 5-20 resistant isolates. This capability addresses a practical limitation of static, manually curated catalogues, whose performance for less well-characterised drugs depends on the accumulation and periodic expert review of sufficient evidence. Clinically, these complementary gains could reduce both unnecessary exclusion of potentially active agents and missed detection of emerging resistance. The ablation and attribution analyses support this interpretation. Removing semantic drug embeddings caused a marked performance loss, indicating that MuseAMR benefits from drug-conditioned genotype-resistance representations, rather than from shared genomic features alone. Importantly, Integrated Gradients attributions remained concentrated on canonical resistance loci such as *katG*, *rpoB*, *embB*, and *gyrA*, demonstrating that the broader whole-genome representation does not disconnect the model from established resistance biology. This interpretability enhances clinical trust and readiness ^13^.

This study has several limitations. First, although MuseAMR simplifies genomic preprocessing, the inherent infrastructure costs and turnaround time of WGS remain restrictive for some decentralised primary care settings. Second, the model relies on binary phenotypic DST results, which introduces label noise and obscures the continuous nature of minimum inhibitory concentrations. Third, while our attribution analysis highlights complex mutational backgrounds potentially associated with epistasis or compensatory evolution, systematic wet-lab validation and perturbation analysis are required to conclusively establish causal mechanisms. Future work should prioritise integrating 3D protein structure mapping and expanding international sample libraries for emerging therapies, further solidifying the framework as a cornerstone for global precision tuberculosis management.

## Methods

### Study design

To bridge the clinical translation gap, we designed a three-tiered validation framework that explicitly separates model development, geographic generalisation assessment, and extension to emerging drugs. (Figure 1) The study was organised into three datasets: a primary development dataset for model training, cross-validation, and hyperparameter tuning (*N* = 10,886 isolates from nine regions or countries ^7^); an independent external validation dataset for geographic generalisation testing (*N* = 18,334 isolates); and a dataset for transfer learning experiments on emerging drugs (*N* = 8,119 isolates).

MuseAMR was trained as a multi-label model to jointly predict resistance to nine anti-tuberculosis drugs. Drug-level outputs were also aggregated into clinically actionable resistance categories aligned with WHO care pathways, including HR-TB, RR-TB, MDR-TB, and pre-XDR-TB. Internal performance was evaluated by cross-validation. External performance was assessed by zero-shot testing across six regional cohorts, followed by local fine-tuning using 10% and 50% of regional data. For the emerging drug extension analysis, we compared transfer learning from the pretrained MuseAMR backbone with models trained from scratch under few-shot settings using *k* = 5 or *k* = 20 resistant isolates matched with susceptible isolates at a 1:5 ratio. Post-hoc feature attribution employed Integrated Gradients ^20^.

This study was a secondary analysis of de-identified, isolate-level MTB WGS data and paired pDST records from the CRyPTIC and from datasets reported in previously published studies. No patients or volunteers were recruited for the present study, no new biological specimens were collected, and the study team did not access names, contact details, clinical records, or other directly identifiable personal information. Ethical approval and participant consent for sample collection, sequencing, and data generation were provided in the original contributing studies, where required. The present analysis used only non-identifiable bacterial genomic and phenotypic resistance data and did not require additional participant consent.

### Datasets

#### Sample sources and inclusion criteria

We utilised MTB isolates from the Comprehensive Resistance Prediction for Tuberculosis: an International Consortium (CRyPTIC) database as the primary dataset ^7^. For external validation, we curated a validation dataset from 25 published global studies providing paired MTB WGS and DST data (Supplementary Table S3). Drug-region combinations with fewer than 200 valid isolates were excluded.

#### Phenotype standardisation and label definition

For the primary dataset, phenotypic susceptibility was determined using 96-well 7H9 broth microdilution (UKMYC5). Binary resistance labels were assigned using established ECOFF/ECV thresholds ^21^. For the external validation dataset, phenotypic records were harmonised following the three-tier protocol of Walker et al. (2021) ^9^: (I) methods and critical concentrations currently endorsed by the WHO (e.g., LJ medium, MGIT); (II) WHO-aligned methods where critical concentrations were either outdated or not explicitly detailed; and (III) quantitative MIC methods without predefined critical concentrations, which we converted into categorical outcomes using plate-specific epidemiological thresholds. Any phenotypic records derived from protocols falling outside these three categories were excluded from our analysis.

#### Genomic preprocessing and input representation

Whole-genome sequencing reads (Illumina) were mapped to the H37Rv reference genome (GenBank NC_00096292.3). *clockwork* (https://github.com/iqbal-lab-org/clockwork) was used for variant calling, and *gnomonicus* (https://github.com/oxfordmmm/gnomonicus) for genetic mutation annotation. For MuseAMR, mutations are formalised into standardised vocabularies (e.g., *KatG*_S315T) that capture the gene name, mutation type, and relative position. Each isolate is represented as a permutation-invariant set of mutations.

For baseline models requiring fixed-length feature vectors, the input space was restricted to variants within the 33 Tier 1 genes defined by the WHO Catalogue (Supplementary Table S5). This aligns with the feature sets used in the source studies we compared against and avoids the curse of dimensionality. The input was structured as a binary occurrence matrix (*N*_samples_ × *n*_uniquemut_), where each entry signifies the presence or absence of a mutation.

### Multimodal Semantic Fusion Model for Antimicrobial Resistance Prediction (MuseAMR)

The MuseAMR consists of four core modules: mutation embedding module, residual convolutional backbone, gating screening, and drug-mutation cross-attention fusion. The model architecture is shown in Figure 6.

**Figure 6.**
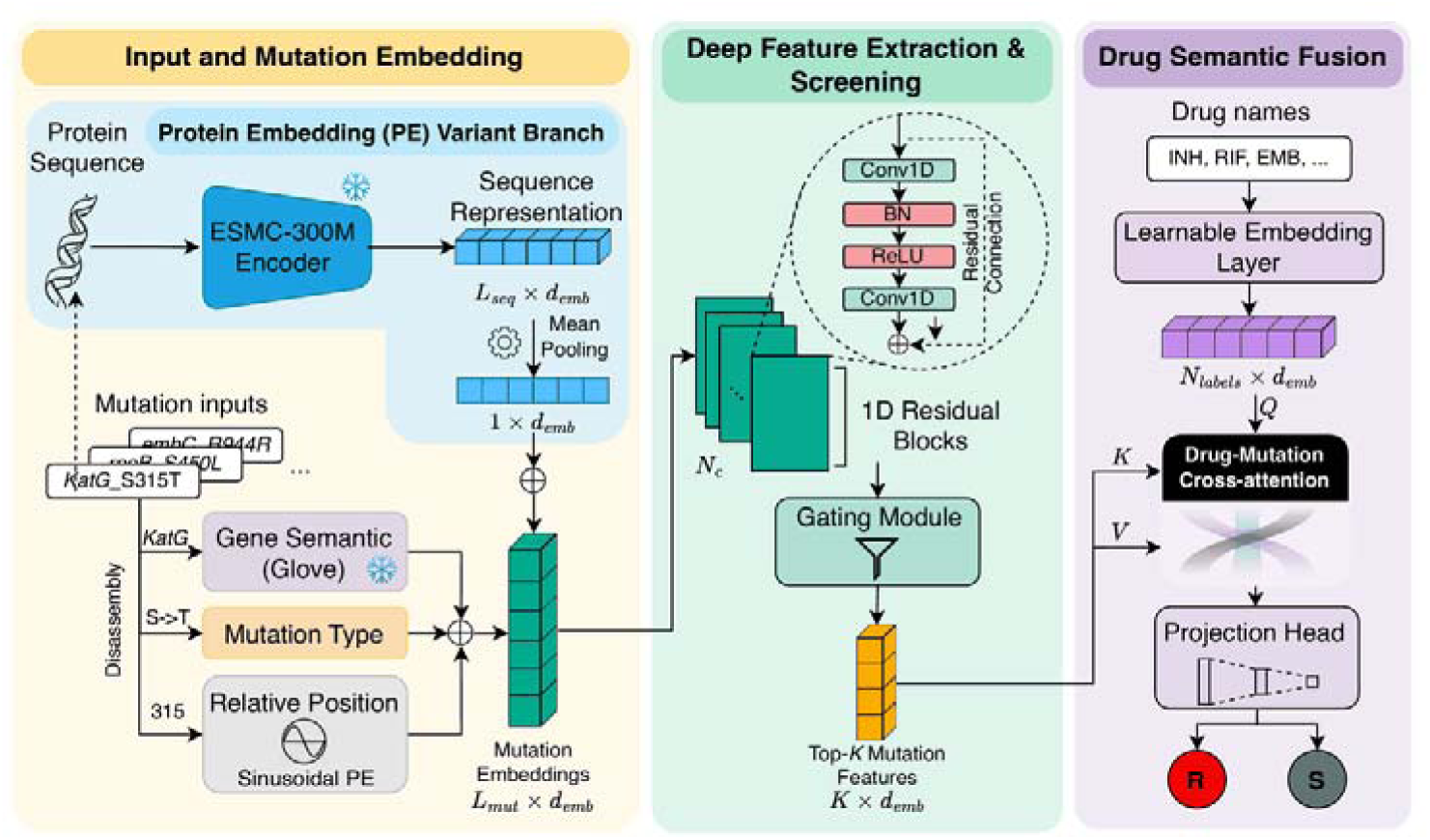
Overview of the unified MuseAMR architecture and its Protein Embedding (PE) variant. The architecture integrates mutation and protein context for downstream prediction. In the base module (bottom), mutation tokens are embedded by integrating gene semantic, mutation type, and positional information, yielding initial mutation embeddings of size *L*_mut_ × *d*_emb_ (where *L*_mut_ is the maximum number of tokens and *d*_emb_ is the embedding dimension). For the MuseAMR (PE) variant (top branch), the original protein sequence corresponding to each mutated gene is encoded in parallel by ESMC-300M. This produces representations of size *L*_seq_ × *d*_emb_, which are mean-pooled into a sequence-level vector of size *d*_emb_ and added (+) to the mutation embeddings to enhance protein context. The fused embeddings are then processed through a stack of *N_c_* 1D residual blocks, followed by a gating module that selects the most informative Top-*K* embeddings (*K* × *d*_emb_). Simultaneously, drug names are mapped into learnable embeddings (*N*_labels_ × *d*_emb_). A cross-attention module then uses these drug embeddings as queries, and the selected mutation embeddings as keys and values, to capture drug-specific genotype--resistance associations. Finally, a projection head predicts resistance (R) or susceptibility (S) for each drug.

#### Mutation semantic embedding

First, in order to fully capture the contextual information of every gene mutation, we decomposed each mutation into three sub-features, including gene semantic information (i.e., the gene in which the mutation is located), mutation type (e.g., SNP, insertion, and deletion), and positional information (i.e., the location of the mutation within the gene).

Taking the *KatG*_S315T mutation as an example, the model parses it into three parts: (1) the gene name (KatG) associated with the mutation is extracted and initialised as a high-dimensional dense vector (d_emb) using GloVe pre-trained embeddings^22^; this step retains global semantic information derived from mutation frequency and co-occurring mutations in each gene; (2) the amino acid substitution pattern (“S-T”) is encoded using a randomly initialised learnable embedding layer and mapped to the same dimensional space; and (3) a sinusoidal positional embedding is introduced for the relative position where the mutation occurs (pos = 315), as follows:

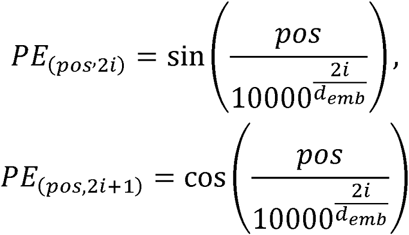

All three embedding vectors are summed along the feature dimension, generating an initial mutation feature matrix with a dimension of *L_mut_* × *d_emb_*, where *L_mut_* is the maximum number of mutations among all samples.

#### Deep feature encoding

After semantic embedding, we designed a feature encoder to extract high-order local correlation features in long sequences while improving computational efficiency. The encoder consists of two components: deep feature extraction and screening.

The deep feature extraction network is composed of *N_c_* one-dimensional residual blocks. Each residual block contains two `Conv1D` layers, each followed by batch normalisation and a ReLU activation function^23^. A residual connection is introduced within the module:

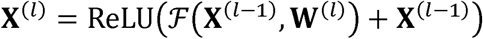

where **X**^(*l*)^ represents the input feature matrix of *l^th^* residual block.

The gated screening mechanism adaptively evaluates the importance of each mutation feature and selects the most representative Top-*K* embedded features, thereby reducing the feature dimension to *K × d_”emb”_*.

#### Semantic-aware drug-mutation fusion and classification head

With the latent mutation embedding, the model receives the target drug name as agent contextual input (such as INH or RIF). These discrete drug labels are mapped into drug representation vectors with dimensions *N_labels_ × d_emb_* through a random initialised embedding layer. Subsequently, the drug embedding serves as the query, while the Top-*K* key features obtained from the previous module are used as keys and values. A cross-attention mechanism is then applied to produce drug-specific vectors with *d_emb_* dimensions. These vectors are finally fed into a unified classification head to yield a binary classification result (resistant [R] / susceptible [S]) for each drug simultaneously.

#### MuseAMR protein embedded variant incorporating protein functional features (MuseAMR (PE))

In complex antibiotic resistance mechanisms, amino acid mutations could contribute to resistance by changing the folding state of the protein or the microenvironment of the drug-binding pocket. Therefore, we designed an extended variant of the basic model, termed MuseAMR (protein embedded, PE), which explicitly accounts for the potential impact of genetic mutations on the three-dimensional structure and function of proteins.

Specifically, MuseAMR (PE) incorporates protein sequence embeddings from ESM Cambrian (ESMC) into the original mutation embeddings. ESMC is a pre-trained large language model for proteins and is the next-generation model following the well-known ESM-2, developed by Meta AI^24,25^. By pre-training on large-scale protein sequence data, the protein sequence representations generated by ESMC can capture the underlying biological information of proteins, including protein structure, evolutionary direction, or other relevant features.

For mutated genes (such as the *KatG* gene), we extracted the complete protein amino acid sequences from the genome and applied the pre-trained ESMC-300M model to obtain a protein representation matrix (dimension *L_seq_ × d_emb_*). This matrix was then average-pooled into a *d_emb_*-dimensional vector, serving as an overall representation of the protein. Finally, this *d_emb_*-dimensional ESMC embedding vector was summed with the mutation embedding features described above. Due to limited computational resources, protein embeddings were generated only for mutations occurring in Tier1 genes as defined by the WHO Catalogue.

### Model training and evaluation

#### Loss function and hyperparameter search

The conventional cross-entropy loss biases the model towards predicting the majority class (S). To better address the severe class imbalance in the dataset, we employed the Asymmetric Loss (ASL) function, which dynamically re-weights positive and negative samples^26^. ASL introduces a probability-shifting mechanism to suppress easy negative samples. Let the true label for a given drug be *y* ε {0,1}, where 1 denotes R and 0 denotes S, and let *p* represent the predicted probability of resistance. The ASL loss is defined as:

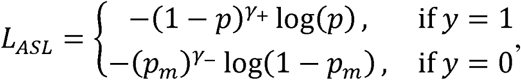

where

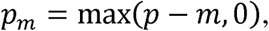

with γ+ and γ- denoting the focusing parameter for positive and negative samples, respectively, and *m* denoting the margin threshold.

By decoupling the focusing parameters for positive and negative samples, ASL applies stronger down-weighting to easily classified negative samples (i.e., the abundant susceptible cases), thereby reducing their dominance during training. In this study, we set γ+ = 0.0, γ- = 4.0, and *m* = 0.05.

The backbone feature embedding dimension (*d_emb_*) of the model was 768. In the MuseAMR (PE) variant that incorporates features from the ESMC protein language model, this dimension was expanded to 960. To constrain model complexity and stabilise gradient updates, the weight decay was set to 0.5, and the global gradient clipping threshold was set to 0.5. Learning rate scheduling was implemented using the `ReduceLROnPlateau` strategy, which adaptively decreases the learning rate when the validation performance reaches a plateau, thereby facilitating stable convergence during the later stages of training. The maximum number of training epochs was set to 100.

During validation, Macro-AUPRC (macro-averaged area under the precision-recall curve) was used for performance monitoring. If the validation Macro-AUPRC failed to exhibit a meaningful improvement for 20 consecutive epochs, training was automatically terminated and the best-performing model weights were retained. In addition, multiple dropout layers were incorporated into the model during training, with a dropout rate set to 0.35.

We employed the `Optuna` framework to optimise key network hyperparameters. The search space included the following parameters: for the convolutional feature extraction layers, the number of 1D residual blocks (*N_c_*) was discretely searched within 2, 4, 6, 8; for the gating module, the number of key mutations selected (K) was discretely explored within 16, 32, 64, 128; and for the cross-feature fusion module, the number of multi-head attention heads (*n*_heads_) was optimised within 4, 8, 16. For the learning rate, the initial learning rate (lr) was continuously sampled within the logarithmic space between 10^-5^ and 10^-3^. The final optimised hyperparameter settings are as follows:

- *N_c_* = 6
- *K* = 32
- *lr* = 5 × 10^-5^
- *n*_heads_ = 8

To enhance the model’s ability to generalise local genomic features and to reduce overfitting to the absolute ordering of mutation tokens, we introduced a data augmentation strategy during training. Specifically, for a subset of training samples, an additional augmented instance was generated by randomly shuffling the order of mutations within the input sequence while preserving the underlying mutation set. In this way, the model was exposed to multiple orderings of the same genomic profile, which increased training diversity without altering the biological content of each sample. Augmented samples were included in training at a ratio of 1:2 relative to the original samples.

#### Evaluation metrics

Given the severe imbalance between resistant and susceptible samples in the drug resistance prediction task, this study adopted the area under the receiver operating characteristic curve (AUROC), area under the precision-recall curve (AUPRC), as well as sensitivity and specificity, as the evaluation metrics. For machine learning methods, the classification threshold was determined by selecting the point on the ROC curve closest to the upper-left corner.

It should be noted that the WHO Catalogue method, which served as the clinical reference benchmark, is fundamentally based on deterministic mutation-matching rules. Therefore, only sensitivity and specificity were reported for this method in comparative analyses, as it does not generate continuous probability outputs or support adjustable decision thresholds.

### Experiments

#### Baseline methods

Three strong baseline methods, including the WHO Catalogue, logistic regression (LR), and wide-deep neural network (WDNN) ^12^, were included to represent rule-based classification, traditional machine learning, and state-of-the-art deep learning, respectively.

The WHO Catalogue, the catalogue of Mycobacterium tuberculosis complex mutations associated with drug resistance released by WHO, is a strong rule-matching method based on expert consensus and large-scale statistics. If the genomic mutations of an MTB isolate contain any mutation marked by the WHO Catalogue as being associated with drug resistance (e.g., *KatG*_S315T corresponds to INH resistance), the isolate is classified as R; otherwise, it is classified as S. Using this as a baseline, the aim was to verify whether our proposed data-driven model (1) exceeded the predictive level of the traditional rule-based method and (2) could capture complex or rare resistance patterns not yet included in the WHO Catalogue. In this study, the WHO Catalogue assessed resistance to eight drugs, and the summary of the catalogue is shown in Supplementary Table S4.

LR is widely used in genotype-phenotype association studies and has been the optimal model for single-drug resistance prediction in many existing studies. To enable multi-label classification tasks, we replaced the traditional way of constructing multiple independent binary classifiers for each drug with an equivalent neural network architecture. Specifically, the LR model is constructed as a Multi-Layer Perceptron (MLP) with only input and output layers, to efficiently predict multi-drug resistance.

WDNN remains a strong model for many drug resistance classification tasks. The WDNN network consists of two parts: wide (equivalent to logistic regression) and deep (MLP network), intended to capture simple linear correlations and nonlinear characteristics, respectively, to achieve a balance between model performance and efficiency.

#### Experimental details

This study comprised the following experiments: (1) a quantitative comparison between the MuseAMR approach and baseline models on the primary development dataset with cross-validation, with the resistance/susceptibility (R/S) labels for nine anti-tuberculosis drugs as prediction targets; the accuracy of model-based clinical resistance categorisation was evaluated afterwards; (2) a region-stratified comparison between MuseAMR and baseline methods on the external validation dataset to assess generalisation performance across different geographic sources; (3) an interpretability analysis of the model using the Integrated Gradients (IG) method; (4) extension of the MuseAMR framework to resistance prediction tasks for emerging anti-tuberculosis drugs through transfer fine-tuning, followed by comparison with models trained from scratch; and (5) ablation study.

For the primary dataset, we performed 5-fold random splitting and conducted cross-validation experiments. For all tasks, performance metrics are reported as the mean and standard deviation across the five folds to evaluate both model performance and stability. Specifically, the data were first divided into training and test sets at a ratio of 4:1. Subsequently, 10% of the training set was further separated as a validation set. This resulted in an effective sample proportion of 72%, 8%, and 20% for the training, validation, and test sets, respectively, in each fold. The overall sample size and the distribution of resistant cases for each drug were maintained consistently across all folds (as shown in Supplementary Table S5).

For the model-based clinical grading analysis, we derived WHO-aligned resistance categories from the predicted resistance outputs for nine drugs. Specifically, cases were classified as: isoniazid-resistant TB (HR-TB) if predicted to be resistant to INH but susceptible to RIF; rifampicin-resistant TB (RR-TB) if predicted to be resistant to RIF; multidrug-resistant TB (MDR-TB) if predicted to be resistant to both INH and RIF; and pre-extensively drug-resistant TB (pre-XDR-TB) if predicted to be resistant to RIF and to at least one fluoroquinolone, which in this study referred to either LEV or MXF.

Because these resistance categories are not mutually exclusive, each category was evaluated as an independent binary classification task. The clinical grading results derived from the model-predicted drug resistance profiles were compared against the grading results determined from the pDST labels, which served as the ground truth, and the metrics were then calculated.

For the external validation experiment, we evaluated each method using the best-performing model checkpoint (except the WHO Catalogue) on the primary dataset to perform inference (or subsequent fine-tuning) on the external validation dataset, thereby obtaining the predicted logit values for each sample. For our model, we further conducted fine-tuning using 10% and 50% of the external validation data to assess its few-shot adaptation capability. During fine-tuning, the embedding, residual blocks, and gating layers were frozen, whereas the cross-attention and classifier layers were left trainable. To estimate the robustness of performance on the external validation dataset, bootstrap resampling was performed 500 times, and all evaluation metrics were reported as the mean values together with their 95% confidence intervals (95% CI).

Model interpretability analysis was conducted using IG as a post hoc attribution method. For an arbitrary input sample *x*, a reference baseline input *x*′, (defined as a unified placeholder input containing no valid mutations), and the logit output *F_c_*(*x*) corresponding to the target drug label *c*, the attribution score of the i-th input feature is defined as:

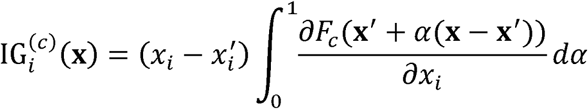

Sample-level IG scores were calculated separately for each drug label. The samples were then stratified into four groups according to prediction outcomes, namely true positives (TP), false positives (FP), true negatives (TN), and false negatives (FN). Within each group, the IG contributions of the same mutation feature were averaged across samples, and the top 20 mutations with the largest absolute attribution values were subsequently identified by ranking the mean absolute contributions.

Few-shot transfer learning experiments for novel drugs were conducted under an episode-based evaluation framework, with `BCEWithLogitsLoss` adopted for the single-label binary classification task. In each episode, the training set comprised a fixed number *k* of resistant samples together with negatively matched susceptible samples at a ratio of 1:5, yielding 5*k* susceptible samples. During transfer, the backbone parameters of the pretrained base model were entirely frozen. The embedding of the target novel drug was initialised as a learnable linear combination of the embedding vectors of the original nine drugs, while the classification head was unfrozen. In this stage, the gating module hyperparameter *K* was set to 128, the learning rate was fixed at 10^-5^, and training was performed for 10 epochs uniformly. Transfer performance was evaluated under the settings of *k* = 5 and *k* = 20.

To validate the contribution of individual modules in MuseAMR, we conducted four ablation experiments: removal of GloVe-based initialization, denoted as NoGlv; removal of semantic label embedding, denoted as NoLabelEmb; removal of positional encoding, denoted as NoPE; and removal of the feature-selection gating module, denoted as NoSP.

#### Hardware and computational details

All model training and evaluation experiments in this study were conducted on a single NVIDIA RTX 4080 Super GPU equipped with 16 GB VRAM. To overcome computational bottlenecks under limited GPU memory and maximise model throughput, we adopted `DeepSpeed`^27^-accelerated Bfloat16 mixed-precision training.

The training batch size was set to 4. To effectively increase the optimisation stability while remaining within the memory constraints, gradient accumulation with a step size of 8 was applied, resulting in an equivalent batch size of 32. Under this combined hardware and engineering setup, the average training time per epoch was approximately 1.5 minute.

### Declaration of generative AI and AI-assisted technologies in the writing process

During the preparation of this work, the author(s) used ChatGPT to improve the language and readability. Following the use of this tool, the author(s) carefully reviewed and edited the content as necessary and take full responsibility for the final version of the manuscript.

## Supplementary information

Please see supplementary file for supplementary tables and figures.

## Acknowledgements

This study was funded by Shanghai three-year (2023-2025) action plan to strengthen the public health system (grant No. GWVI-11.1-05). The funders of the study had no role in the study design, data collection, data analysis, data interpretation, or writing of the report. The computations in this paper were run on the Siyuan-1 cluster supported by the Center for High Performance Computing at Shanghai Jiao Tong University.

## Data availability

This study used de-identified bacterial isolate-level genomic and phenotypic data from public sources, and no new individual participant data were collected. The CRyPTIC dataset is publicly available through the Bacterial and Viral Bioinformatics Resource Center ^7,28^. Accession information and references for the literature-derived external validation datasets are provided in the Supplementary Material. The curated isolate list, data dictionary, and derived model inputs and outputs are available from the corresponding author on reasonable request after publication, subject to the data-use restrictions of the original sources.

## Code availability

The source code for data preprocessing, model training, and evaluation is available from the corresponding author on reasonable request after publication.

## Author contributions

C.L., H.W. and Y.Y. conceived and designed the study. C.L. developed the MuseAMR framework, performed model training, validation and statistical analyses, and drafted the manuscript. H.Z. curated the external validation datasets and contributed to data harmonisation. X.W. contributed to genomic data preprocessing and optimisation of the bioinformatics workflow. Y.L. contributed to interpretation of the findings and critical revision of the manuscript. Y.Y. contributed to study design, model experiments, data interpretation and manuscript revision. H.W. supervised the overall study. C.L., H.Z. and Y.Y. directly accessed and verified the underlying data. All authors reviewed and approved the final manuscript.

