## Supplementary material for "Robust AI Framework for Comprehensive Tuberculosis Drug Resistance Profiling with Rapid Adaptability"

June 5, 2026

#### Contents

**1 Source of samples in primary dataset and external validation dataset. 2**

**2 Regional distribution of samples and drug resistance profiles in the external validation dataset. 4**

**3 WHO catalogues of resistance-associated genes and mutations for antibiotics 6**

**4 Sample distribution across training, validation, and test sets for each drug 7**

**5 Quantitative comparison of MuseAMR and baselines in the primary dataset 8**

**6 External validation 9**

**7 Model interpretations 15**

**8 Ablation study 16**

### 1 Source of samples in primary dataset and external validation dataset.

**Table S1: Geographical distribution of samples in the primary dataset.** Sample counts and corresponding proportions are shown for each contributing country, with countries further classified by continent. The proportion for each country was calculated relative to the total number of samples in the primary dataset.

| Country | Continent | Sample $N$ | Proportion (%) |
|---|---|---|---|
| Peru | South America | 2637 | 24.22 |
| South Africa | Africa | 1998 | 18.35 |
| Italy | Europe | 1333 | 12.25 |
| India | Asia | 1311 | 12.04 |
| Vietnam | Asia | 1264 | 11.61 |
| Germany | Europe | 1089 | 10.00 |
| China | Asia | 916 | 8.41 |
| Brazil | South America | 333 | 3.06 |
| Sweden | Europe | 5 | 0.05 |

**Table S2: Geographical distribution of samples in the external validation dataset.** Sample counts and corresponding proportions are shown for each contributing region or country. The proportion for each region was calculated relative to the total number of samples in the external validation dataset.

| Region | Sample $N$ | Proportion (%) |
|---|---|---|
| Africa | 3165 | 17.26 |
| America | 1638 | 8.93 |
| Asia (except China) | 1752 | 9.56 |
| China | 8650 | 47.18 |
| Europe | 1367 | 7.46 |
| Oceania | 1762 | 9.61 |

**Table S3: Summary of the published studies contributing to the literature-derived external validation datasets used in this study.** The table summarises the basic characteristics of the included studies, including study type, country or region, and sample size.

| Study | Study type | Region (Country) | Sample size |
|---|---|---|---|
| Mejía-Ponce PM et al. (2023) [1] | surveillance | Thailand | 276 |
| Noroc E et al. (2023) [2] | surveillance | Moldova | 268 |
| Vasiliauskaitė L et al. (2024) [3] | method evaluation | Poland, Lithuania | 208 |
| Vīksna A et al. (2023) [4] | method evaluation | Latvia | 63 |
| Che Y et al. (2022) [5] | method evaluation | China | 59 |
| Guimarães AEDS et al. (2021) [6] | method evaluation | Brazil | 71 |
| Morey-León G et al. (2023) [7] | method evaluation | Ecuador | 88 |
| Qu, J. et al. (2024) [8] | method development | China | 82 |
| Thorpe, J. et al. (2024) [9] | method evaluation | Thailand | 59 |
| Mok, S. et al. (2021) [10] | method evaluation | Ireland | 147 |
| Finci, Iris et al. (2022) [11] | epidemiological analysis | Mixed | 900 |
| Liu, Kuang-Hung et al. (2024) [12] | epidemiological analysis | China (Taiwan) | 297 |
| Zhang, Xiaomei et al. (2024) [13] | epidemiological analysis | Australia | 1812 |
| Pei, Shaojun et al. (2024) [14] | surveillance | China | 8170 |

Continued on next page

| Study | Study type | Region (Country) | Sample size |
|---|---|---|---|
| Kim D et al. (2024) [15] | method development | – | 5463 |
| Che Y et al. (2024) [16] | epidemiological analysis | China | 130 |
| Van Nguyen, H. et al. (2024) [17] | genomic analysis | Vietnam | 233 |
| Lundeberg, E.E. et al. (2023) [18] | – | – | 110 |
| Merker, M. et al. (2021) [19] | epidemiological analysis | Cameroon | 195 |
| Chaiyachat, Pratchakan et al. (2021) [20] | surveillance | Thailand | 276 |
| Billard-Pomares, Typhaine et al. (2022) [21] | surveillance | France | 227 |
| Macedo, Rita et al. (2023) [22] | genomic analysis | Portugal | 100 |
| Olivenca, Francisco et al. (2022) [23] | genomic analysis | Portugal | 172 |
| Bateson, Anna et al. (2022) [24] | – | – | 330 |
| Avika Dixit et al. (2024) [25] | epidemiological analysis | Mixed | 12,023 |

#### 2 Regional distribution of samples and drug resistance profiles in the external validation dataset.

**Table S4: Sample sizes and resistance proportions of individual drugs across source regions in the external validation dataset.** The table summarises, for each geographic region, the numbers of resistant and susceptible isolates, the total number of isolates with available phenotypic labels, and the corresponding resistance ratio for each anti-tuberculosis drug. Resistance ratio was calculated as the percentage of resistant isolates among all isolates with valid drug susceptibility results for the corresponding drug within each region.

| Region | Drug | Resistant | Susceptible | Total | Resistance ratio (%) |
|---|---|---|---|---|---|
| Africa | INH | 635 | 2526 | 3161 | 20.09 |
| | RIF | 502 | 2660 | 3162 | 15.88 |
| | EMB | 235 | 579 | 814 | 28.87 |
| | LEV | 58 | 64 | 122 | 47.54 |
| | MXF | 195 | 1627 | 1822 | 10.70 |
| | AMI | 203 | 1610 | 1813 | 11.20 |
| | KAN | 264 | 1753 | 2017 | 13.09 |
| | ETH | 85 | 110 | 195 | 43.59 |
| America | INH | 379 | 1167 | 1546 | 24.51 |
| | RIF | 309 | 1318 | 1627 | 18.99 |
| | EMB | 217 | 1392 | 1609 | 13.49 |
| | KAN | 184 | 82 | 266 | 69.17 |
| Asia (except China) | INH | 795 | 813 | 1608 | 49.44 |
| | RIF | 753 | 861 | 1614 | 46.65 |
| | EMB | 258 | 323 | 581 | 44.41 |
| | LEV | 98 | 364 | 462 | 21.21 |
| | MXF | 93 | 1237 | 1330 | 6.99 |
| | AMI | 31 | 588 | 619 | 5.01 |
| | KAN | 63 | 806 | 869 | 7.25 |
| | ETH | 86 | 192 | 278 | 30.94 |
| China | INH | 1270 | 7370 | 8640 | 14.70 |
| | RIF | 704 | 7945 | 8649 | 8.14 |
| | EMB | 231 | 8419 | 8650 | 2.67 |
| | LEV | 443 | 8204 | 8647 | 5.12 |
| | MXF | 380 | 8269 | 8649 | 4.39 |
| | AMI | 107 | 8542 | 8649 | 1.24 |
| | KAN | 539 | 8107 | 8646 | 6.23 |
| | ETH | 314 | 8335 | 8649 | 3.63 |
| Europe | INH | 459 | 630 | 1089 | 42.15 |
| | RIF | 435 | 925 | 1360 | 31.99 |
| | EMB | 185 | 720 | 905 | 20.44 |
| | LEV | 19 | 313 | 332 | 5.72 |
| | MXF | 24 | 210 | 234 | 10.26 |
| | AMI | 46 | 458 | 504 | 9.13 |
| | KAN | 165 | 293 | 458 | 36.03 |
| | ETH | 37 | 213 | 250 | 14.80 |
| Oceania | INH | 205 | 1551 | 1756 | 11.67 |
| | RIF | 80 | 1682 | 1762 | 4.54 |

Continued on next page

**Table S4 (Continued)**

| Region | Drug | Resistant | Susceptible | Total | Resistance ratio (%) |
|---|---|---|---|---|---|
| | EMB | 39 | 1723 | 1762 | 2.21 |
| Total | INH | 3743 | 14 057 | 17 800 | 21.03 |
| | RIF | 2783 | 15 391 | 18 174 | 15.31 |
| | EMB | 1165 | 13 156 | 14 321 | 8.13 |
| | LEV | 623 | 9025 | 9648 | 6.46 |
| | MXF | 697 | 11 590 | 12 287 | 5.67 |
| | AMI | 392 | 11 289 | 11 681 | 3.36 |
| | KAN | 1215 | 11 047 | 12 262 | 9.91 |
| | ETH | 522 | 8854 | 9376 | 5.57 |

### 3 WHO catalogues of resistance-associated genes and mutations for antibiotics

**Table S5: List of Tier 1 genes related to MTB resistance identified by WHO.** The table summarises the genes classified by the WHO as Tier 1 resistance-associated genes for each anti-tuberculosis drug, comparing the older version with the 2021 and 2023 WHO catalogues. Tier 1 genes represent genes with well-established evidence linking genetic variation to phenotypic drug resistance and are commonly used as core targets in molecular resistance prediction and mutation screening.

| Drug | Tier 1 Genes (old) | Tier1 genes (2021) | Tier1 genes (2023) |
|---|---|---|---|
| AMI (Amikacin) | *rrs, eis, whiB7* | *rrs, eis* | *rrs, eis* |
| BDQ (Bedaquiline) | *pepQ, Rv0678, mmpL5, mmpS5, atpE* | – | *atpE, pepQ, Rv0678* |
| CFZ (Clofazimine) | *pepQ, Rv0678, mmpL5, mmpS5* | – | *pepQ, Rv0678* |
| DLM (Delamanid) | *fgd1, ddn, fbiA, fbiB, fbiC, Rv2983* | *ddn* | *ddn, fbiA, fbiB, fbiC, fgd1, Rv2983* |
| EMB (Ethambutol) | *embA, embB, embC* | *embA, embB* | *embB* |
| ETH (Ethionamide) | *inhA, ethA* | *inhA, ethA* | *inhA, ethA* |
| INH (Isoniazid) | *ahpC, inhA, katG* | *inhA, katG* | *inhA, katG* |
| KAN (Kanamycin) | *rrs, eis, whiB7* | *rrs, eis* | *rrs, eis* |
| LEV (Levofloxacin) | – | *gyrA, gyrB* | *gyrA, gyrB* |
| LZD (Linezolid) | *rplC, rrl* | *rplC* | *rplC, rrl* |
| MXF (Moxifloxacin) | *gyrA, gyrB* | *gyrA, gyrB* | *gyrA, gyrB* |
| RIF (Rifampicin) | *rpoB* | *rpoB* | *rpoB* |
| PZA (Pyrazinamide) | *pncA, clpC1, panD* | *pncA* | *pncA* |
| STM (Streptomycin) | *rrs, rpsL, gid, whiB7, Rv1258c* | *rrs, rpsL, gid* | *rrs, rpsL, gid* |
| CAP (Capreomycin) | *rrs, tlyA* | *rrs, tlyA* | *rrs, tlyA* |

**Table S6: WHO catalogue of resistance-associated mutations for 8 anti-tuberculosis drugs.** The table summarises the resistance-associated mutation entries curated by WHO for eight anti-tuberculosis drugs, including the treatment line, the genes involved, the total number of mutation entries, and representative mutations for each drug.

| Drug | Lines | Gene (n) | Total | Representative mutation(s) |
|---|---|---|---|---|
| INH | First-line | *katG* (135), *inhA* (8) | 143 | *katG_S315T*, *katG_S315N*, *katG_S315I*, *katG_S315R*, *inhA_g-154a*, *inhA_S94A* |
| RIF | First-line | *rpoB* (136) | 136 | *rpoB_S450L*, *rpoB_H445D*, *rpoB_H445Y*, *rpoB_H445R*, *rpoB_D435V*, *rpoB_D435Y* |
| EMB | First-line | *embB* (13) | 13 | *embB_M306V*, *embB_M306I*, *embB_M306L*, *embB_G406A*, *embB_G406D* |
| AMI | Second-line | *rrs* (3), *eis* (1) | 4 | *rrs_a1401g*, *rrs_c1402t*, *rrs_g1484t*, *eis_c-14t* |
| KAN | Second-line | *eis* (5), *rrs* (3) | 8 | *rrs_a1401g*, *eis_c-14t*, *eis_g-10a*, *eis_c-12t* |
| LEV | Second-line | *gyrA* (10), *gyrB* (8) | 18 | *gyrA_D94G*, *gyrA_D94A*, *gyrA_D94N*, *gyrA_A90V*, *gyrA_S91P* |
| MXF | Second-line | *gyrA* (10), *gyrB* (8) | 18 | *gyrA_D94G*, *gyrA_D94A*, *gyrA_D94N*, *gyrA_A90V*, *gyrA_S91P* |
| ETH | Second-line | *ethA* (279), *inhA* (8) | 287 | *ethA_deletion*, *ethA_LoF*, *ethA_t-7c*, *inhA_g-154a*, *inhA_S94A* |

### 4 Sample distribution across training, validation, and test sets for each drug

**Table S7: Distribution of resistant (R) and sensitive (S) samples for each drug training, validation, and test splits.** The table summarises the numbers of resistant and susceptible isolates and the corresponding total sample size in the training, validation, and test sets for each drug.

| Drug | Train | | | Validation | | | Test | | |
|---|---|---|---|---|---|---|---|---|---|
| | R | S | Total | R | S | Total | R | S | Total |
| INH | 3216 | 3627 | 6843 | 358 | 403 | 761 | 893 | 1008 | 1901 |
| RIF | 2482 | 3956 | 6438 | 276 | 440 | 716 | 690 | 1099 | 1789 |
| EMB | 1050 | 4345 | 5395 | 117 | 483 | 600 | 292 | 1207 | 1499 |
| LEV | 849 | 4740 | 5589 | 94 | 528 | 622 | 236 | 1317 | 1553 |
| MXF | 714 | 4164 | 4878 | 79 | 463 | 542 | 198 | 1157 | 1355 |
| AMI | 430 | 6022 | 6452 | 48 | 669 | 717 | 120 | 1673 | 1793 |
| KAN | 547 | 6161 | 6708 | 61 | 685 | 746 | 152 | 1712 | 1864 |
| ETH | 856 | 5485 | 6341 | 95 | 610 | 705 | 238 | 1524 | 1762 |
| RFB | 2300 | 4918 | 7218 | 256 | 547 | 803 | 639 | 1367 | 2006 |

### 5 Quantitative comparison of MuseAMR and baselines in the primary dataset

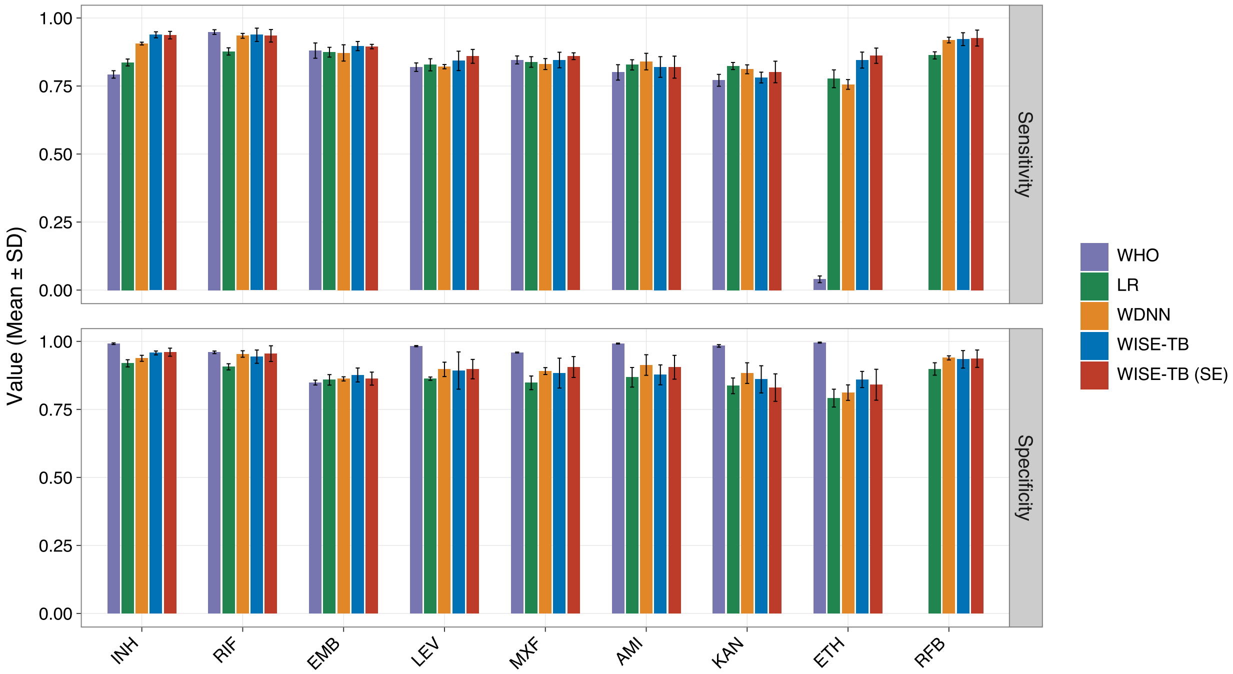


Figure S1: **Comparison of drug-specific sensitivities and specificities between MuseAMR and baseline models.** The figure shows the predictive performance of MuseAMR and baseline models in terms of sensitivity and specificity by drugs. Each bar represents the mean performance value calculated across cross-validation folds, and the error bars indicate the corresponding standard deviation, reflecting the variability of model performance.

### 6 External validation

**Table S8: Detailed external validation results of all models by source region and drug.** The table reports the performance of MuseAMR and the baseline models for each drug within each geographic region in the external validation analysis. For every region–drug–model combination, AUROC, AUPRC, sensitivity, and specificity were presented. Since the WHO Catalogue method does not generate continuous prediction scores, AUROC and AUPRC are not applicable and are therefore shown as "–". Bold values indicate the best-performing model for a given metric within each region and drug.

| Region | Model | Drug | AUROC | AUPRC | Sensitivity | Specificity |
|---|---|---|---|---|---|---|
| Africa | WHO | INH | – | – | 0.690 | **0.997** |
| Africa | LR | INH | 0.896 | 0.846 | 0.759 | 0.967 |
| Africa | WDNN | INH | 0.936 | 0.902 | 0.838 | 0.971 |
| Africa | MuseAMR | INH | 0.932 | 0.887 | 0.820 | 0.970 |
| Africa | MuseAMR (10% ) | INH | 0.945 | 0.910 | 0.871 | 0.978 |
| Africa | MuseAMR (50% ) | INH | **0.946** | **0.916** | **0.879** | 0.977 |
| Africa | WHO | RIF | – | – | 0.952 | **0.985** |
| Africa | LR | RIF | 0.953 | 0.904 | 0.890 | 0.967 |
| Africa | WDNN | RIF | 0.977 | 0.929 | 0.926 | 0.971 |
| Africa | MuseAMR | RIF | 0.980 | 0.928 | **0.964** | 0.975 |
| Africa | MuseAMR (10% ) | RIF | **0.982** | 0.930 | 0.960 | 0.982 |
| Africa | MuseAMR (50% ) | RIF | 0.978 | **0.944** | 0.952 | 0.978 |
| Africa | WHO | EMB | – | – | **0.957** | 0.813 |
| Africa | LR | EMB | 0.889 | 0.680 | 0.830 | 0.796 |
| Africa | WDNN | EMB | 0.908 | 0.764 | 0.902 | 0.789 |
| Africa | MuseAMR | EMB | 0.905 | 0.748 | **0.957** | 0.789 |
| Africa | MuseAMR (10% ) | EMB | **0.938** | **0.821** | 0.921 | 0.845 |
| Africa | MuseAMR (50% ) | EMB | **0.938** | 0.763 | 0.933 | **0.862** |
| Africa | WHO | MXF | – | – | 0.810 | **0.980** |
| Africa | LR | MXF | 0.976 | 0.850 | 0.944 | 0.942 |
| Africa | WDNN | MXF | 0.977 | 0.795 | 0.959 | 0.945 |
| Africa | MuseAMR | MXF | 0.932 | 0.667 | 0.882 | 0.887 |
| Africa | MuseAMR (10% ) | MXF | 0.977 | 0.802 | 0.953 | 0.926 |
| Africa | MuseAMR (50% ) | MXF | **0.984** | **0.884** | **0.978** | 0.935 |
| Africa | WHO | AMI | – | – | 0.916 | **0.999** |
| Africa | LR | AMI | 0.968 | 0.909 | 0.911 | 0.960 |
| Africa | WDNN | AMI | 0.950 | 0.881 | 0.926 | 0.978 |
| Africa | MuseAMR | AMI | 0.976 | 0.827 | 0.916 | 0.991 |
| Africa | MuseAMR (10% ) | AMI | 0.981 | 0.960 | 0.934 | 0.986 |
| Africa | MuseAMR (50% ) | AMI | **0.988** | **0.967** | **0.950** | 0.974 |
| Africa | WHO | KAN | – | – | 0.852 | **0.997** |
| Africa | LR | KAN | 0.950 | 0.874 | **0.917** | 0.897 |
| Africa | WDNN | KAN | 0.901 | 0.810 | 0.860 | 0.731 |
| Africa | MuseAMR | KAN | 0.949 | 0.798 | 0.845 | 0.986 |
| Africa | MuseAMR (10% ) | KAN | **0.960** | **0.921** | 0.869 | 0.981 |

Continued on next page

Table S8 (continued)

| Region | Model | Drug | AUROC | AUPRC | Sensitivity | Specificity |
|---|---|---|---|---|---|---|
| Africa | MuseAMR (50% ) | KAN | 0.957 | 0.907 | 0.868 | 0.976 |
| America | WHO | INH | – | – | 0.850 | **0.999** |
| America | LR | INH | 0.897 | 0.882 | 0.794 | 0.969 |
| America | WDNN | INH | 0.951 | 0.936 | 0.892 | 0.981 |
| America | MuseAMR | INH | **0.967** | 0.950 | 0.894 | 0.989 |
| America | MuseAMR (10% ) | INH | 0.963 | 0.953 | 0.916 | 0.985 |
| America | MuseAMR (50% ) | INH | 0.964 | **0.958** | **0.917** | 0.988 |
| America | WHO | RIF | – | – | 0.916 | **0.995** |
| America | LR | RIF | 0.931 | 0.872 | 0.841 | 0.933 |
| America | WDNN | RIF | 0.971 | 0.943 | 0.900 | 0.963 |
| America | MuseAMR | RIF | **0.981** | 0.950 | **0.961** | 0.938 |
| America | MuseAMR (10% ) | RIF | 0.976 | **0.955** | 0.935 | 0.978 |
| America | MuseAMR (50% ) | RIF | 0.968 | 0.914 | 0.929 | 0.979 |
| America | WHO | EMB | – | – | **0.954** | **0.985** |
| America | LR | EMB | 0.959 | 0.774 | 0.124 | 0.983 |
| America | WDNN | EMB | 0.966 | 0.873 | 0.917 | 0.981 |
| America | MuseAMR | EMB | 0.949 | 0.677 | 0.908 | 0.960 |
| America | MuseAMR (10% ) | EMB | 0.971 | 0.950 | 0.943 | 0.968 |
| America | MuseAMR (50% ) | EMB | **0.974** | **0.958** | 0.939 | 0.977 |
| America | WHO | KAN | – | – | 0.967 | 0.988 |
| America | LR | KAN | 0.965 | 0.980 | 0.984 | 0.732 |
| America | WDNN | KAN | **0.995** | **0.998** | **0.989** | 0.854 |
| America | MuseAMR | KAN | 0.988 | 0.995 | 0.967 | **1.000** |
| America | MuseAMR (10% ) | KAN | 0.986 | 0.994 | 0.963 | **1.000** |
| America | MuseAMR (50% ) | KAN | 0.987 | 0.996 | 0.959 | **1.000** |
| Asia (except China) | WHO | INH | – | – | 0.732 | **0.990** |
| Asia (except China) | LR | INH | 0.925 | 0.938 | 0.755 | 0.948 |
| Asia (except China) | WDNN | INH | 0.969 | 0.976 | 0.877 | 0.968 |
| Asia (except China) | MuseAMR | INH | 0.954 | 0.961 | 0.836 | 0.980 |
| Asia (except China) | MuseAMR (10% ) | INH | 0.971 | 0.976 | 0.929 | 0.963 |
| Asia (except China) | MuseAMR (50% ) | INH | **0.982** | **0.985** | **0.947** | 0.964 |
| Asia (except China) | WHO | RIF | – | – | 0.930 | **0.972** |
| Asia (except China) | LR | RIF | 0.898 | 0.897 | 0.761 | 0.889 |
| Asia (except China) | WDNN | RIF | 0.935 | 0.935 | 0.843 | 0.943 |
| Asia (except China) | MuseAMR | RIF | 0.967 | 0.966 | 0.927 | 0.965 |
| Asia (except China) | MuseAMR (10% ) | RIF | 0.961 | 0.955 | 0.930 | 0.965 |
| Asia (except China) | MuseAMR (50% ) | RIF | **0.976** | **0.971** | **0.949** | 0.970 |
| Asia (except China) | WHO | EMB | – | – | **0.810** | **0.796** |
| Asia (except China) | LR | EMB | 0.761 | 0.703 | 0.663 | 0.693 |
| Asia (except China) | WDNN | EMB | 0.773 | 0.694 | 0.736 | 0.709 |
| Asia (except China) | MuseAMR | EMB | 0.799 | 0.754 | 0.802 | 0.663 |
| Asia (except China) | MuseAMR (10% ) | EMB | 0.832 | 0.791 | 0.784 | 0.749 |

Continued on next page

Table S8 (continued)

| Region | Model | Drug | AUROC | AUPRC | Sensitivity | Specificity |
|---|---|---|---|---|---|---|
| Asia (except China) | MuseAMR (50% ) | EMB | **0.836** | **0.807** | 0.797 | 0.773 |
| Asia (except China) | WHO | LEV | – | – | 0.847 | **0.876** |
| Asia (except China) | LR | LEV | 0.717 | 0.447 | **0.888** | 0.250 |
| Asia (except China) | WDNN | LEV | 0.824 | 0.622 | 0.816 | 0.706 |
| Asia (except China) | MuseAMR | LEV | 0.724 | 0.535 | 0.622 | 0.808 |
| Asia (except China) | MuseAMR (10% ) | LEV | 0.840 | 0.680 | 0.782 | 0.872 |
| Asia (except China) | MuseAMR (50% ) | LEV | **0.883** | **0.767** | 0.864 | 0.832 |
| Asia (except China) | WHO | MXF | – | – | 0.882 | **0.935** |
| Asia (except China) | LR | MXF | **0.909** | 0.447 | **0.925** | 0.723 |
| Asia (except China) | WDNN | MXF | 0.893 | 0.530 | 0.774 | 0.868 |
| Asia (except China) | MuseAMR | MXF | 0.845 | 0.465 | 0.742 | 0.889 |
| Asia (except China) | MuseAMR (10% ) | MXF | 0.868 | 0.469 | 0.738 | 0.897 |
| Asia (except China) | MuseAMR (50% ) | MXF | 0.899 | **0.616** | 0.814 | 0.904 |
| Asia (except China) | WHO | AMI | – | – | **0.903** | 0.991 |
| Asia (except China) | LR | AMI | 0.767 | 0.276 | 0.774 | 0.626 |
| Asia (except China) | WDNN | AMI | 0.901 | 0.688 | 0.677 | 0.944 |
| Asia (except China) | MuseAMR | AMI | 0.877 | 0.722 | 0.742 | 0.988 |
| Asia (except China) | MuseAMR (10% ) | AMI | 0.888 | 0.757 | 0.741 | **0.998** |
| Asia (except China) | MuseAMR (50% ) | AMI | **0.942** | **0.813** | 0.833 | 0.996 |
| Asia (except China) | WHO | KAN | – | – | **0.778** | **0.988** |
| Asia (except China) | LR | KAN | 0.762 | 0.404 | 0.730 | 0.593 |
| Asia (except China) | WDNN | KAN | **0.827** | **0.562** | 0.635 | 0.867 |
| Asia (except China) | MuseAMR | KAN | 0.695 | 0.494 | 0.460 | 0.954 |
| Asia (except China) | MuseAMR (10% ) | KAN | 0.758 | 0.499 | 0.527 | 0.891 |
| Asia (except China) | MuseAMR (50% ) | KAN | 0.794 | 0.500 | 0.731 | 0.767 |
| China | WHO | INH | – | – | 0.577 | **0.996** |
| China | LR | INH | 0.788 | 0.644 | 0.611 | 0.940 |
| China | WDNN | INH | 0.863 | 0.734 | 0.677 | 0.968 |
| China | MuseAMR | INH | 0.812 | 0.617 | 0.649 | 0.909 |
| China | MuseAMR (10% ) | INH | **0.882** | 0.780 | 0.751 | 0.948 |
| China | MuseAMR (50% ) | INH | 0.876 | **0.781** | **0.755** | 0.957 |
| China | WHO | RIF | – | – | 0.807 | **0.993** |
| China | LR | RIF | 0.861 | 0.683 | 0.709 | 0.937 |
| China | WDNN | RIF | 0.891 | 0.739 | 0.784 | 0.937 |
| China | MuseAMR | RIF | 0.841 | 0.556 | 0.699 | 0.900 |
| China | MuseAMR (10% ) | RIF | **0.927** | **0.835** | **0.833** | 0.983 |
| China | MuseAMR (50% ) | RIF | 0.922 | 0.818 | 0.809 | 0.989 |
| China | WHO | EMB | – | – | 0.732 | **0.985** |
| China | LR | EMB | 0.876 | 0.452 | 0.615 | 0.975 |
| China | WDNN | EMB | 0.881 | 0.421 | 0.667 | 0.974 |
| China | MuseAMR | EMB | 0.820 | 0.349 | 0.790 | 0.808 |
| China | MuseAMR (10% ) | EMB | 0.897 | **0.522** | **0.792** | 0.944 |

Continued on next page

Table S8 (continued)

| Region | Model | Drug | AUROC | AUPRC | Sensitivity | Specificity |
|---|---|---|---|---|---|---|
| China | MuseAMR (50% ) | EMB | **0.902** | 0.508 | 0.762 | 0.976 |
| China | WHO | LEV | – | – | 0.555 | **0.997** |
| China | LR | LEV | 0.733 | 0.280 | 0.411 | 0.939 |
| China | WDNN | LEV | 0.733 | 0.381 | 0.420 | 0.969 |
| China | MuseAMR | LEV | 0.645 | 0.129 | 0.419 | 0.841 |
| China | MuseAMR (10% ) | LEV | 0.776 | 0.307 | 0.556 | 0.933 |
| China | MuseAMR (50% ) | LEV | **0.789** | **0.463** | **0.623** | 0.914 |
| China | WHO | MXF | – | – | 0.571 | **0.994** |
| China | LR | MXF | 0.744 | 0.258 | 0.511 | 0.861 |
| China | WDNN | MXF | 0.750 | 0.361 | 0.497 | 0.953 |
| China | MuseAMR | MXF | 0.656 | 0.154 | 0.454 | 0.829 |
| China | MuseAMR (10% ) | MXF | 0.781 | 0.260 | **0.658** | 0.809 |
| China | MuseAMR (50% ) | MXF | **0.795** | **0.448** | 0.598 | 0.944 |
| China | WHO | AMI | – | – | 0.346 | **0.999** |
| China | LR | AMI | 0.687 | 0.115 | 0.308 | 0.950 |
| China | WDNN | AMI | 0.712 | 0.274 | 0.383 | 0.966 |
| China | MuseAMR | AMI | 0.599 | 0.065 | **0.449** | 0.759 |
| China | MuseAMR (10% ) | AMI | 0.674 | 0.231 | 0.327 | 0.990 |
| China | MuseAMR (50% ) | AMI | **0.715** | **0.326** | 0.345 | **0.999** |
| China | WHO | KAN | – | – | 0.085 | **1.000** |
| China | LR | KAN | 0.350 | 0.072 | 0.124 | 0.854 |
| China | WDNN | KAN | 0.430 | 0.122 | 0.212 | 0.868 |
| China | MuseAMR | KAN | 0.524 | 0.093 | 0.347 | 0.799 |
| China | MuseAMR (10% ) | KAN | 0.612 | 0.130 | 0.607 | 0.576 |
| China | MuseAMR (50% ) | KAN | **0.792** | **0.228** | **0.745** | 0.756 |
| China | WHO | ETH | – | – | 0.025 | **0.999** |
| China | LR | ETH | 0.648 | 0.109 | 0.172 | 0.958 |
| China | WDNN | ETH | 0.631 | 0.107 | 0.178 | 0.960 |
| China | MuseAMR | ETH | 0.684 | 0.163 | **0.548** | 0.787 |
| China | MuseAMR (10% ) | ETH | 0.688 | 0.102 | 0.451 | 0.847 |
| China | MuseAMR (50% ) | ETH | **0.707** | **0.240** | 0.464 | 0.864 |
| Europe | WHO | INH | – | – | 0.850 | **0.983** |
| Europe | LR | INH | 0.934 | 0.939 | 0.850 | 0.943 |
| Europe | WDNN | INH | 0.947 | **0.952** | **0.891** | 0.921 |
| Europe | MuseAMR | INH | 0.934 | 0.936 | 0.858 | 0.973 |
| Europe | MuseAMR (10% ) | INH | 0.941 | 0.935 | 0.880 | 0.977 |
| Europe | MuseAMR (50% ) | INH | **0.949** | 0.949 | 0.880 | 0.975 |
| Europe | WHO | RIF | – | – | 0.903 | 0.970 |
| Europe | LR | RIF | 0.943 | 0.919 | 0.878 | 0.929 |
| Europe | WDNN | RIF | 0.952 | 0.929 | 0.913 | 0.943 |
| Europe | MuseAMR | RIF | 0.947 | 0.940 | **0.924** | 0.973 |
| Europe | MuseAMR (10% ) | RIF | 0.955 | 0.942 | 0.923 | 0.980 |

Continued on next page

Table S8 (continued)

| Region | Model | Drug | AUROC | AUPRC | Sensitivity | Specificity |
|---|---|---|---|---|---|---|
| Europe | MuseAMR (50% ) | RIF | **0.969** | **0.963** | 0.919 | **0.989** |
| Europe | WHO | EMB | – | – | 0.708 | 0.929 |
| Europe | LR | EMB | **0.905** | 0.735 | 0.573 | **0.958** |
| Europe | WDNN | EMB | 0.902 | **0.740** | 0.714 | 0.876 |
| Europe | MuseAMR | EMB | 0.844 | 0.577 | **0.892** | 0.731 |
| Europe | MuseAMR (10% ) | EMB | 0.893 | 0.724 | 0.849 | 0.848 |
| Europe | MuseAMR (50% ) | EMB | 0.904 | 0.738 | 0.796 | 0.930 |
| Europe | WHO | LEV | – | – | **0.895** | **0.994** |
| Europe | LR | LEV | 0.723 | 0.317 | 0.842 | 0.419 |
| Europe | WDNN | LEV | 0.843 | 0.462 | 0.842 | 0.805 |
| Europe | MuseAMR | LEV | 0.826 | 0.260 | **0.895** | 0.719 |
| Europe | MuseAMR (10% ) | LEV | **0.871** | 0.218 | 0.875 | 0.819 |
| Europe | MuseAMR (50% ) | LEV | 0.868 | **0.523** | 0.778 | 0.973 |
| Europe | WHO | MXF | – | – | 0.583 | **0.995** |
| Europe | LR | MXF | 0.582 | 0.199 | **0.750** | 0.100 |
| Europe | WDNN | MXF | 0.622 | 0.280 | 0.667 | 0.605 |
| Europe | MuseAMR | MXF | **0.633** | **0.455** | 0.375 | 0.976 |
| Europe | MuseAMR (10% ) | MXF | 0.602 | 0.424 | 0.364 | 0.984 |
| Europe | MuseAMR (50% ) | MXF | 0.631 | 0.379 | 0.583 | 0.847 |
| Europe | WHO | AMI | – | – | 0.609 | **0.978** |
| Europe | LR | AMI | 0.872 | 0.630 | 0.739 | 0.803 |
| Europe | WDNN | AMI | 0.770 | 0.619 | 0.609 | 0.904 |
| Europe | MuseAMR | AMI | 0.881 | 0.455 | 0.739 | 0.897 |
| Europe | MuseAMR (10% ) | AMI | **0.893** | **0.674** | 0.829 | 0.914 |
| Europe | MuseAMR (50% ) | AMI | 0.877 | 0.569 | **0.842** | 0.892 |
| Europe | WHO | KAN | – | – | 0.879 | **0.942** |
| Europe | LR | KAN | **0.947** | **0.897** | **0.982** | 0.666 |
| Europe | WDNN | KAN | 0.881 | 0.869 | 0.824 | 0.805 |
| Europe | MuseAMR | KAN | 0.853 | 0.645 | 0.848 | 0.782 |
| Europe | MuseAMR (10% ) | KAN | 0.923 | 0.816 | 0.857 | 0.886 |
| Europe | MuseAMR (50% ) | KAN | 0.930 | 0.851 | 0.824 | 0.907 |
| Europe | WHO | ETH | – | – | 0.216 | **0.991** |
| Europe | LR | ETH | 0.857 | 0.463 | 0.865 | 0.765 |
| Europe | WDNN | ETH | 0.874 | 0.561 | 0.811 | 0.751 |
| Europe | MuseAMR | ETH | 0.887 | 0.471 | 0.892 | 0.812 |
| Europe | MuseAMR (10% ) | ETH | 0.875 | 0.517 | **0.943** | 0.688 |
| Europe | MuseAMR (50% ) | ETH | **0.899** | **0.687** | 0.870 | 0.820 |
| Oceania | WHO | INH | – | – | 0.659 | **0.999** |
| Oceania | LR | INH | 0.801 | 0.619 | 0.556 | 0.941 |
| Oceania | WDNN | INH | 0.907 | 0.806 | 0.761 | 0.974 |
| Oceania | MuseAMR | INH | 0.933 | 0.833 | 0.790 | 0.974 |
| Oceania | MuseAMR (10% ) | INH | 0.930 | 0.827 | **0.818** | 0.966 |

Continued on next page

Table S8 (continued)

| Region | Model | Drug | AUROC | AUPRC | Sensitivity | Specificity |
|---|---|---|---|---|---|---|
| Oceania | MuseAMR (50% ) | INH | **0.934** | **0.856** | 0.813 | 0.988 |
| Oceania | WHO | RIF | – | – | 0.887 | **0.997** |
| Oceania | LR | RIF | 0.922 | 0.725 | 0.850 | 0.900 |
| Oceania | WDNN | RIF | 0.940 | 0.779 | 0.863 | 0.956 |
| Oceania | MuseAMR | RIF | 0.978 | **0.936** | 0.963 | 0.992 |
| Oceania | MuseAMR (10% ) | RIF | 0.981 | 0.918 | 0.972 | 0.984 |
| Oceania | MuseAMR (50% ) | RIF | **0.992** | 0.876 | **0.974** | 0.980 |
| Oceania | WHO | EMB | – | – | 0.923 | **0.989** |
| Oceania | LR | EMB | 0.895 | 0.555 | 0.641 | 0.984 |
| Oceania | WDNN | EMB | 0.941 | 0.589 | 0.795 | 0.987 |
| Oceania | MuseAMR | EMB | 0.975 | 0.493 | 0.949 | 0.950 |
| Oceania | MuseAMR (10% ) | EMB | 0.987 | 0.678 | 0.944 | 0.984 |
| Oceania | MuseAMR (50% ) | EMB | **0.994** | **0.756** | **1.000** | 0.976 |

### 7 Model interpretations

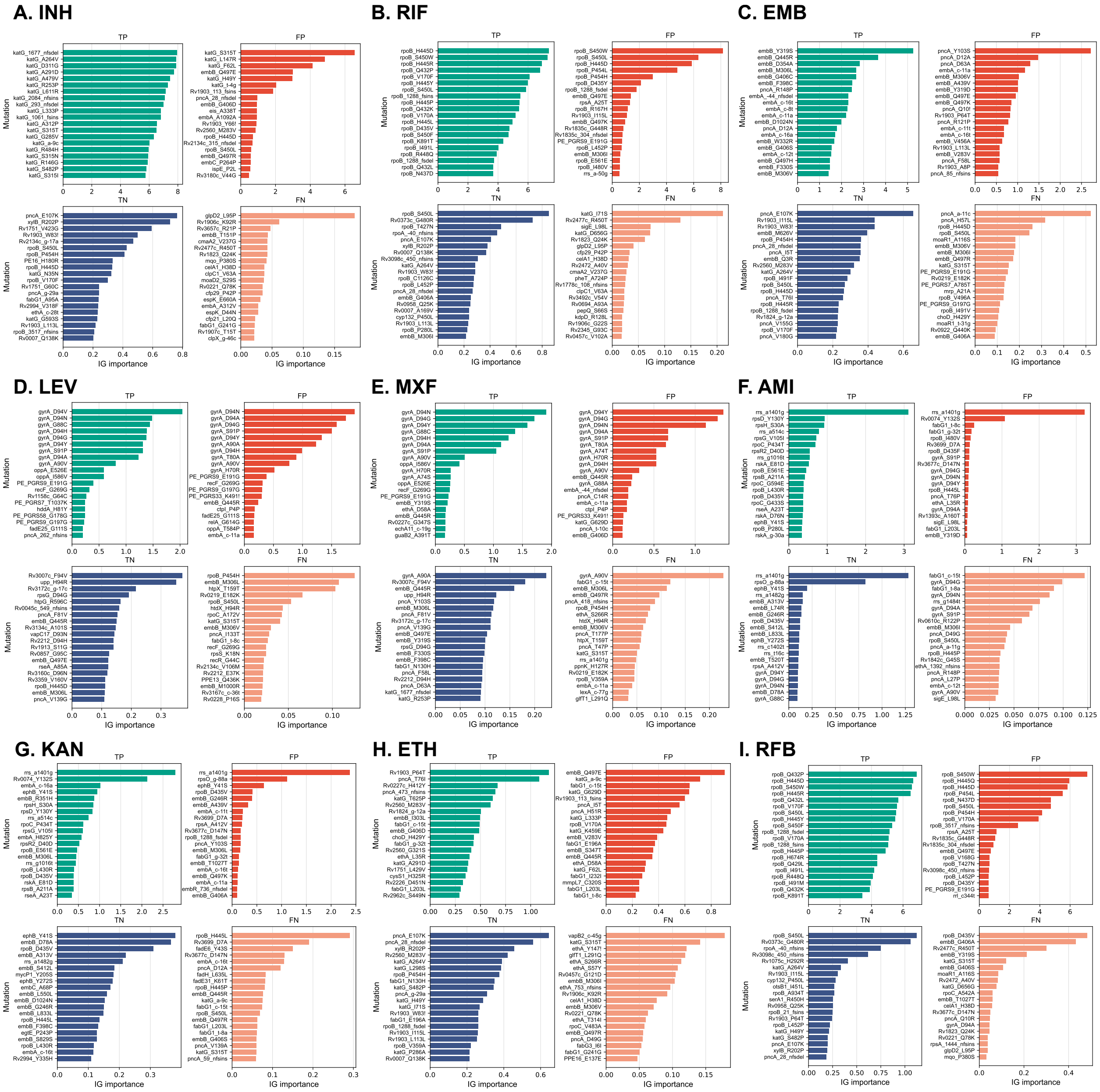


Figure S2: **Top 20 mutation importance (IG) values for the four prediction groups of each drug.** For each drug, the top 20 mutations with the highest absolute integrated gradients (IG) importance are displayed separately for true positives (TP), false positives (FP), true negatives (TN), and false negatives (FN). Panels show results for (A) INH, (B) RIF, (C) EMB, (D) LEV, (E) MXF, (F) AMI, (G) KAN, (H) ETH, and (I) RFB. Bar length indicates the mean IG importance of each mutation within the corresponding prediction group, with higher values reflecting greater contribution to the model prediction. Colors denote TP (green), FP (red), TN (blue), and FN (orange).

### 8 Ablation study

To evaluate the contribution of individual modules, we conducted four ablation experiments: removal of GloVe-based initialization (*NoGlv*); removal of semantic label embedding (*NoLabelEmb*); removal of positional encoding (*NoPE*); and removal of the feature-selection gating module (*NoSP*). The results are reported in two parts: aggregated performance for first-line and second-line drugs (Table S9), and drug-specific performance (S10).

**Table S9:** Results of the ablation study on first- and second-line drugs.

| Ablation type | Drug | Metric | Full model result | Ablation result |
|---|---|---|---|---|
| NoGlv | First-line | AUPRC | **0.908** ± **0.012** | 0.906 ± 0.013 |
| NoGlv | First-line | AUROC | 0.960 ± 0.005 | **0.961** ± **0.005** |
| NoGlv | Second-line | AUPRC | 0.768 ± 0.073 | **0.793** ± **0.046** |
| NoGlv | Second-line | AUROC | 0.921 ± 0.017 | **0.927** ± **0.015** |
| NoLabelEmb | First-line | AUPRC | **0.908** ± **0.012** | 0.868 ± 0.012 |
| NoLabelEmb | First-line | AUROC | **0.960** ± **0.005** | 0.941 ± 0.009 |
| NoLabelEmb | Second-line | AUPRC | **0.768** ± **0.073** | 0.757 ± 0.048 |
| NoLabelEmb | Second-line | AUROC | **0.921** ± **0.017** | 0.911 ± 0.017 |
| NoPE | First-line | AUPRC | **0.908** ± **0.012** | 0.870 ± 0.015 |
| NoPE | First-line | AUROC | **0.960** ± **0.005** | 0.941 ± 0.008 |
| NoPE | Second-line | AUPRC | **0.768** ± **0.073** | 0.726 ± 0.062 |
| NoPE | Second-line | AUROC | **0.921** ± **0.017** | 0.909 ± 0.014 |
| NoSP | First-line | AUPRC | **0.908** ± **0.012** | 0.902 ± 0.013 |
| NoSP | First-line | AUROC | **0.960** ± **0.005** | 0.958 ± 0.004 |
| NoSP | Second-line | AUPRC | 0.768 ± 0.073 | **0.813** ± **0.035** |
| NoSP | Second-line | AUROC | 0.921 ± 0.017 | **0.937** ± **0.011** |

In contrast to the full MuseAMR model, removal of semantic label embedding or positional encoding resulted in a consistent decline in predictive performance. Specifically, the model without semantic label embedding showed an average decrease of 0.040 in AUPRC and 0.019 in AUROC for first-line drug prediction tasks, and decreases of 0.011 and 0.010, respectively, for second-line drug prediction tasks. Similarly, after removing positional encoding, the average AUPRC and AUROC for first-line drugs decreased by 0.038 and 0.019, respectively, while for second-line drugs the corresponding decreases were 0.042 and 0.012. These findings indicate that both semantic label embedding and positional encoding are core contributing modules, with the removal of positional encoding exerting a particularly pronounced impact on second-line AUPRC.

In contrast, removing GloVe initialization or the gating-based feature selection module did not lead to a consistent deterioration in performance relative to the full MuseAMR model. For example, both ablated variants showed performance gains on second-line drug prediction tasks. After removing GloVe initialization, the mean AUPRC for second-line drugs increased from 0.768 to 0.793 ($\Delta = +0.025$), and the mean AUROC increased from 0.921 to 0.927 ($\Delta = +0.006$). Likewise, removal of the gating mechanism improved second-line AUPRC and AUROC by 0.045 and 0.016, respectively. For first-line drug prediction tasks, however, neither ablation showed substantial performance differences compared with the full model.

**Table S10:** The results of ablation study on each drug

| Ablation type | Drug | Metric | Full model result | Ablation result |
|---|---|---|---|---|
| NoGlv | AMI | AUPRC | 0.711 ± 0.142 | **0.799** ± **0.056** |
| NoGlv | AMI | AUROC | 0.909 ± 0.021 | **0.924** ± **0.018** |
| NoGlv | EMB | AUPRC | **0.802** ± **0.038** | 0.801 ± 0.034 |
| NoGlv | EMB | AUROC | **0.939** ± **0.012** | **0.939** ± **0.010** |

Continued on next page

**Table S10 (continued)**

| Ablation type | Drug | Metric | Full model result | Ablation result |
|---|---|---|---|---|
| NoGlv | ETH | AUPRC | **0.730 ± 0.044** | 0.727 ± 0.084 |
| NoGlv | ETH | AUROC | **0.920 ± 0.016** | 0.917 ± 0.031 |
| NoGlv | INH | AUPRC | **0.970 ± 0.005** | 0.964 ± 0.005 |
| NoGlv | INH | AUROC | **0.972 ± 0.005** | 0.970 ± 0.005 |
| NoGlv | KAN | AUPRC | 0.681 ± 0.111 | **0.749 ± 0.058** |
| NoGlv | KAN | AUROC | 0.896 ± 0.032 | **0.906 ± 0.027** |
| NoGlv | LEV | AUPRC | **0.779 ± 0.109** | 0.776 ± 0.074 |
| NoGlv | LEV | AUROC | **0.918 ± 0.029** | **0.918 ± 0.021** |
| NoGlv | MXF | AUPRC | **0.770 ± 0.082** | 0.761 ± 0.066 |
| NoGlv | MXF | AUROC | 0.919 ± 0.022 | **0.923 ± 0.018** |
| NoGlv | RFB | AUPRC | 0.935 ± 0.021 | **0.943 ± 0.011** |
| NoGlv | RFB | AUROC | 0.966 ± 0.013 | **0.971 ± 0.003** |
| NoGlv | RIF | AUPRC | 0.952 ± 0.011 | **0.954 ± 0.006** |
| NoGlv | RIF | AUROC | 0.970 ± 0.011 | **0.974 ± 0.002** |
| NoGlv | macro | AUPRC | 0.814 ± 0.048 | **0.831 ± 0.032** |
| NoGlv | macro | AUROC | 0.934 ± 0.013 | **0.938 ± 0.012** |
| NoGlv | micro | AUPRC | 0.906 ± 0.021 | **0.910 ± 0.011** |
| NoGlv | micro | AUROC | 0.956 ± 0.010 | **0.958 ± 0.007** |
| NoLabelEmb | AMI | AUPRC | **0.711 ± 0.142** | 0.705 ± 0.114 |
| NoLabelEmb | AMI | AUROC | **0.909 ± 0.021** | 0.898 ± 0.039 |
| NoLabelEmb | EMB | AUPRC | **0.802 ± 0.038** | 0.717 ± 0.041 |
| NoLabelEmb | EMB | AUROC | **0.939 ± 0.012** | 0.918 ± 0.010 |
| NoLabelEmb | ETH | AUPRC | **0.730 ± 0.044** | 0.658 ± 0.045 |
| NoLabelEmb | ETH | AUROC | **0.920 ± 0.016** | 0.879 ± 0.028 |
| NoLabelEmb | INH | AUPRC | **0.970 ± 0.005** | 0.951 ± 0.007 |
| NoLabelEmb | INH | AUROC | **0.972 ± 0.005** | 0.950 ± 0.011 |
| NoLabelEmb | KAN | AUPRC | **0.681 ± 0.111** | 0.668 ± 0.099 |
| NoLabelEmb | KAN | AUROC | **0.896 ± 0.032** | 0.884 ± 0.020 |
| NoLabelEmb | LEV | AUPRC | 0.779 ± 0.109 | **0.807 ± 0.027** |
| NoLabelEmb | LEV | AUROC | 0.918 ± 0.029 | **0.926 ± 0.017** |
| NoLabelEmb | MXF | AUPRC | 0.770 ± 0.082 | **0.793 ± 0.043** |
| NoLabelEmb | MXF | AUROC | 0.919 ± 0.022 | **0.929 ± 0.011** |
| NoLabelEmb | RFB | AUPRC | **0.935 ± 0.021** | 0.913 ± 0.023 |
| NoLabelEmb | RFB | AUROC | **0.966 ± 0.013** | 0.948 ± 0.018 |
| NoLabelEmb | RIF | AUPRC | **0.952 ± 0.011** | 0.937 ± 0.015 |
| NoLabelEmb | RIF | AUROC | **0.970 ± 0.011** | 0.954 ± 0.015 |
| NoLabelEmb | macro | AUPRC | **0.814 ± 0.048** | 0.794 ± 0.032 |
| NoLabelEmb | macro | AUROC | **0.934 ± 0.013** | 0.921 ± 0.014 |
| NoLabelEmb | micro | AUPRC | **0.906 ± 0.021** | 0.885 ± 0.019 |
| NoLabelEmb | micro | AUROC | **0.956 ± 0.010** | 0.943 ± 0.011 |
| NoPE | AMI | AUPRC | **0.711 ± 0.142** | 0.638 ± 0.153 |
| NoPE | AMI | AUROC | **0.909 ± 0.021** | 0.900 ± 0.027 |
| NoPE | EMB | AUPRC | **0.802 ± 0.038** | 0.731 ± 0.025 |
| NoPE | EMB | AUROC | **0.939 ± 0.012** | 0.919 ± 0.006 |
| NoPE | ETH | AUPRC | **0.730 ± 0.044** | 0.636 ± 0.114 |
| NoPE | ETH | AUROC | **0.920 ± 0.016** | 0.871 ± 0.036 |
| NoPE | INH | AUPRC | **0.970 ± 0.005** | 0.942 ± 0.015 |
| NoPE | INH | AUROC | **0.972 ± 0.005** | 0.946 ± 0.013 |
| NoPE | KAN | AUPRC | **0.681 ± 0.111** | 0.613 ± 0.132 |

Continued on next page

**Table S10 (continued)**

| Ablation type | Drug | Metric | Full model result | Ablation result |
|---|---|---|---|---|
| NoPE | KAN | AUROC | **0.896** ± **0.032** | 0.887 ± 0.020 |
| NoPE | LEV | AUPRC | 0.779 ± 0.109 | **0.784** ± **0.094** |
| NoPE | LEV | AUROC | 0.918 ± 0.029 | **0.923** ± **0.027** |
| NoPE | MXF | AUPRC | **0.770** ± **0.082** | **0.770** ± **0.095** |
| NoPE | MXF | AUROC | **0.919** ± **0.022** | **0.919** ± **0.030** |
| NoPE | RFB | AUPRC | **0.935** ± **0.021** | 0.913 ± 0.040 |
| NoPE | RFB | AUROC | **0.966** ± **0.013** | 0.954 ± 0.027 |
| NoPE | RIF | AUPRC | **0.952** ± **0.011** | 0.936 ± 0.025 |
| NoPE | RIF | AUROC | **0.970** ± **0.011** | 0.960 ± 0.023 |
| NoPE | macro | AUPRC | **0.814** ± **0.048** | 0.774 ± 0.042 |
| NoPE | macro | AUROC | **0.934** ± **0.013** | 0.920 ± 0.009 |
| NoPE | micro | AUPRC | **0.906** ± **0.021** | 0.878 ± 0.015 |
| NoPE | micro | AUROC | **0.956** ± **0.010** | 0.944 ± 0.006 |
| NoSP | AMI | AUPRC | 0.711 ± 0.142 | **0.797** ± **0.058** |
| NoSP | AMI | AUROC | 0.909 ± 0.021 | **0.931** ± **0.015** |
| NoSP | EMB | AUPRC | **0.802** ± **0.038** | 0.790 ± 0.028 |
| NoSP | EMB | AUROC | **0.939** ± **0.012** | 0.933 ± 0.009 |
| NoSP | ETH | AUPRC | 0.730 ± 0.044 | **0.747** ± **0.052** |
| NoSP | ETH | AUROC | 0.920 ± 0.016 | **0.928** ± **0.020** |
| NoSP | INH | AUPRC | **0.970** ± **0.005** | 0.963 ± 0.009 |
| NoSP | INH | AUROC | **0.972** ± **0.005** | 0.969 ± 0.004 |
| NoSP | KAN | AUPRC | 0.681 ± 0.111 | **0.745** ± **0.067** |
| NoSP | KAN | AUROC | 0.896 ± 0.032 | **0.914** ± **0.018** |
| NoSP | LEV | AUPRC | 0.779 ± 0.109 | **0.837** ± **0.023** |
| NoSP | LEV | AUROC | 0.918 ± 0.029 | **0.939** ± **0.014** |
| NoSP | MXF | AUPRC | 0.770 ± 0.082 | **0.809** ± **0.036** |
| NoSP | MXF | AUROC | 0.919 ± 0.022 | **0.937** ± **0.017** |
| NoSP | RFB | AUPRC | 0.935 ± 0.021 | **0.944** ± **0.013** |
| NoSP | RFB | AUROC | 0.966 ± 0.013 | **0.971** ± **0.005** |
| NoSP | RIF | AUPRC | **0.952** ± **0.011** | **0.954** ± **0.006** |
| NoSP | RIF | AUROC | 0.970 ± 0.011 | **0.973** ± **0.003** |
| NoSP | macro | AUPRC | 0.814 ± 0.048 | **0.843** ± **0.027** |
| NoSP | macro | AUROC | 0.934 ± 0.013 | **0.944** ± **0.008** |
| NoSP | micro | AUPRC | 0.906 ± 0.021 | **0.913** ± **0.014** |
| NoSP | micro | AUROC | 0.956 ± 0.010 | **0.958** ± **0.004** |
